# Gut microbiota affects prostate cancer risk through steroid hormone biosynthesis

**DOI:** 10.1101/2021.08.19.21262274

**Authors:** Sofia Kalinen, Teemu Kallonen, Marianne Gunell, Otto Ettala, Ivan Jambor, Juha Knaapila, Kari T. Syvänen, Pekka Taimen, Matti Poutanen, Claes Ohlsson, Hannu J. Aronen, Helena Ollila, Sami Pietilä, Laura L. Elo, Tarja Lamminen, Antti J. Hakanen, Eveliina Munukka, Peter J. Boström, the Multi-IMPROD Study group

## Abstract

**Purpose:** Although prostate cancer is the most common cancer in men in Western countries, there is significant variability in geographical incidence. This might result from genetic factors, discrepancies in screening policies or differences in lifestyle. Gut microbiota has been recently associated with cancer progression, but its role in prostate cancer is unclear.

**Methods:** In a prospective multicenter clinical trial (NCT02241122), the gut microbiota profiles of 181 men with a clinical suspicion of prostate cancer were assessed utilizing 16S rRNA gene sequencing. Sequences were assigned to operational taxonomic units, and differential abundance analysis, α- and β-diversities, and predictive functional (PICRUSt) analyses were performed. Additionally, plasma steroid hormone levels were correlated with the predicted microbiota functions.

**Results:** Several differences in the gut microbiota between the subjects with and without prostate cancer were noted. *Prevotella 9*, members of the *Erysipelotrichaceae* family and *Escherichia-Shigella were higher, and Jonquetella, Moryella, Anaeroglobus, Corynebacterium* and CAG-352 were lower in the cancer group. Predictive functional analyses revealed higher 5-α-reductase, copper absorption, and retinal metabolism in the prostate cancer associated microbiome. Plasma testosterone associated negatively with the microbial 5-α-reductase activity (p=0.030). In a subgroup of men taking 5-α-reductase inhibitors (n=17), plasma estrone (p=0.027) and estradiol (p=0.059) levels were lower in men with predicted elevation of the microbial 5-α-reductase function.

**Conclusions:** Gut microbiota of the prostate cancer patients differed significantly compared to benign subjects. Microbial 5-α-reductase, copper absorption and retinol metabolism are potential mechanisms of action. These findings could explain the observed association of lifestyle, geography, and prostate cancer incidence.

## INTRODUCTION

Prostate cancer (PCa), despite its high incidence, has undiscovered details of etiology and pathogenesis.^1,2^ PCa is known to be highly heritable, but also lifestyle, socioeconomic, and environmental factors may affect PCa incidence.^3^ Diet is one of the most widely studied life-style factors, and various nutrients and food products have been reported to be associated with either lower or higher PCa risk.^3,4^

PCa incidence differs markedly between geographical locations, being lowest in Asia and the highest in the Western lifestyle countries.^2^ Differences in ethnicity, genetics, as well as healthcare-related factors such as intensity of prostate-specific antigen (PSA) screening have a significant effect on the reported variability of global PCa incidence.^3^ However, other explanatory factors might also exist. It is of great interest that a high rate of incidental PCa has also been widely reported in the geographical areas of low clinical PCa prevalence.^5^ Additionally, studies conducted in immigrant populations suggest that non-genetic, individual lifestyle factors may significantly affect PCa risk.^6^ Based on these observations, we might expect that prostate carcinogenesis affects a significant portion of aging men, if not all of them. We might also expect that individual lifestyle and environmental factors may either stimulate or inhibit the neoplastic process in the prostate, accounting for the geographical differences observed in clinical PCa. However, the mechanisms mediating how lifestyle affects PCa risk remains unclear.

Gut microbiota (GM), i.e. a collection of all microbes in the gastrointestinal tract, is considered to affect many metabolic pathways and pathogenetic processes in the human body.^7^ Furthermore, gut dysbiosis (disequilibrium of the microbiota) leading to low-grade inflammation has been linked to many cancers, also in organs distant to the intestines.^7^ Chronic inflammation, production of superoxide radicals, growth factors and bacterial genotoxins have all been proposed as mechanisms of action.^7^ Furthermore, the alterations in GM could result from the lifestyles related to lower or higher PCa risk.

A majority of the PCa investigations covering microbiological aspects have studied either prostate tissue or urinary tract microbiota with conflicting results.^8^ To date, the effect of GM on prostate carcinogenesis is poorly documented though there is some evidence of an association with PCa.^8,9^ The association of fecal microbiota with PCa has received little attention, and most studies have very small sample sizes.^8^ Larger studies to date reported differences in GM composition between PCa and non-PCa cases as well as between high-risk and benign-low-risk PCa.^9^ Microbiome analyses also suggested mechanisms of action, including altered folate and arginine metabolism, as well as short-chain fatty acids and IGF-1 signaling.^9^

To assess the fecal microbiota profiles of PCa patients compared to their benign counterparts, we conducted a substudy within a prospective clinical trial (NCT02241122) where microbiological swab samples from men with suspected PCa were 16S sequenced, and a predictive functional analysis (PICRUSt) was performed.^10^ To our knowledge, this is the largest and most detailed clinical trial studying the GM of prostate cancer patients.

## METHODS

### TRIAL DESIGN

The study cohort has been previously reported in detail.^11^ Between February 2015 and March 2017, men with a clinical suspicion of PCa were enrolled at four Finnish hospitals for a prospective, investigator-initiated, open-label, non-randomized trial investigating MRI and biomarkers in PCa diagnosis (Clinicaltrials.gov, NCT02241122). Informed consent was obtained from all study subjects. The study protocol, patient information sheet and informed consent forms were approved by the Ethics Committee of the Hospital District of Southwest Finland (No 6/2014). Ethical principles of the Declaration of Helsinki guiding physicians and medical research involving human subjects (59^th^ World Medical Association General Assembly, Seoul, Korea, 2008) were followed.

### PATIENTS

All men included were clinically suspected of having PCa with PSA ranging from 2.5 to 20.0 μg/l, and/or an abnormal finding in digital rectal examination (DRE). Exclusion criteria included previous prostate biopsy, previous prostate surgery, previous diagnosis of PCa, acute prostatitis, or contraindications for MRI-imaging.

### ASSESSMENTS

After MRI, systematic 12-core prostate and two targeted biopsies from up to two lesions suspected at MRI (Likert score 3-5) were collected from all subjects. Prophylactic antibiotics were given according to the institutional guidelines and have been previously reported in detail.^12^ No enema was administered prior to biopsy. Immediately prior to transrectal biopsies, microbiome samples were collected utilizing sterile rectal swabs (Copan FLOQSwab™, Copan diagnostics Inc., Murrieta, CA, USA) and immediately stored to −20°C. Besides clinical specimens, a detailed questionnaire including family history of PCa, general medical, travel, and smoking history was completed prior to biopsy. Blood samples were collected for steroid assays (plasma).

### MICROBIOLOGICAL SAMPLE PREPARATION FOR 16S RNA SEQUENCING

Stool samples were diluted in 1 ml of sterile saline after which 500 μl were used for the DNA extraction with Semi-Automatic GXT Stool Extraction Kit VER 2.0 and GenoXtract unit (Hain Lifescience GmbH, Nehren, Germany). DNA concentrations were measured with Qubit dsDNA HS Assay Kit and Qubit 2.0 fluorometer (Life Technologies, Carlsbad, USA). Extracted DNAs were stored in −80°C until used. Bacterial V4 gene regions of 16S rRNA were sequenced in three batches with MiSeq® (Illumina, San Diego, California, USA), where negative and positive controls were accompanied.

### DATA ANALYSIS AND BIOINFORMATICS

Statistical analyses were performed with the IBM Statistical Package for the Social Sciences (SPSS®) for Windows, version 26, 64-bit (IBM Corp., Armonk, NY, USA) and R version 4.0.3. Continuous variables were summarized with median and quartiles. The Kolmogorov-Smirnov test evaluated data normality. PCa cases were grouped by two means, each utilizing the International Society of Urological Pathology (ISUP) grade, namely benign cases versus all cancer cases, and benign cases versus ISUP grade group 1, 2-3, and 4-5 cancers, respectively.^13^ Factors potentially impacting either cancer status or GM composition, such as age, body mass index (BMI), recent use of antibiotics, inflammatory bowel diseases (IBD), recent high risk travel history and smoking, were also analyzed with Wilcoxon rank sum, Kruskal-Wallis, Pearson Chi square or Fisher’s exact tests.

Microbial analyses were performed with CLC Genomics Workbench Microbial Genomics module v. 12 (QIAGEN Digital Insights, Aarhus, Denmark). Sequences were assigned to operational taxonomic units (OTU) according to CLC Microbial genomics module workflow. Quality and ambiguous trims were performed with default settings, with the minimum number of nucleotides set to 150. SILVA 16S v132 97% was used as the reference database.^14,15^ α-diversity measures Chao1 and Shannon describing overall bacterial profiles were calculated to evaluate community richness, diversity and evenness with the 10527-rarefaction level. The β-diversity measure, Bray-Curtis dissimilarity, was calculated and the PERMANOVA test with 99999 permutations was used to calculate the p-values. The sequencing batch effect was controlled with β-diversity measures.

Differential abundance analysis with a generalized linear model that applies a negative binomial distribution was also performed. The sequencing batch and the previously mentioned factors potentially impacting gut microbiota were corrected in the analysis. Age was not considered in the analysis because of collinearity. The results were filtered with combined abundance in all samples of more than 100 and prevalence of 10 per cent or more. P-values were corrected using the false discovery rate (FDR) approach. The statistical significance limit was set to a p-value <0.05.

Predictive analysis of functional bacterial genes was constructed utilizing PICRUSt v.1.0 (Phylogenetic Investigation of Communities by Reconstruction of Unobserved States) tool.^10^ PICRUSt was performed with Qiime v. 1.9 to create KEGG orthologs and pathways with the reference data of Greengenes v.13.8.^16–18^ As microbiota and predicted gene products were not normally distributed, data was analyzed with nonparametric Wilcoxon rank sum and Kruskal-Wallis tests and described with medians and 95% confidence intervals.

### PLASMA STEROIDS ASSAY

The steroid analyses of plasma samples were conducted using a LC-MS/MS according to an established method.^19^ Measured steroids included androstenedione, dehydroepiandrosterone (DHEA), dihydrotestosterone (DHT), estradiol, estrone, progesterone, 17-α-hydroxyprogesterone (17-α-OHP), and testosterone (T). In addition, the DHT/T ratio was calculated for the statistical analyses. Wilcoxon rank sum tests and Spearman correlations were used to investigate the association of PICRUSt predicted microbial steroid hormone biosynthesis with plasma steroid status. For Wilcoxon rank sum tests, steroid hormone biosynthesis was divided to high and low according to the median.

## RESULTS

### STUDY SUBJECTS

A total of 364 men entered the trial (see flowchart in **Figure 1**). After excluding patients withdrawing consent (n=24), having MRI artifacts at the time of biopsy (n=2), stool material in rectal swabs insufficient for analyses (n=149), and whose samples failed laboratory quality standards (n=8) (**Supplementary Figure S1**), a total of 181 men were included in the analyses, of whom 167 provided complete questionnaires. Plasma samples were available from 169 cases of which 4 were excluded due to the unknown status of 5-α-reductase inhibitor (5-ARI) medication. Plasma steroids of 5-ARI users were analyzed separately (n=17, 10%).

**Figure 1.**
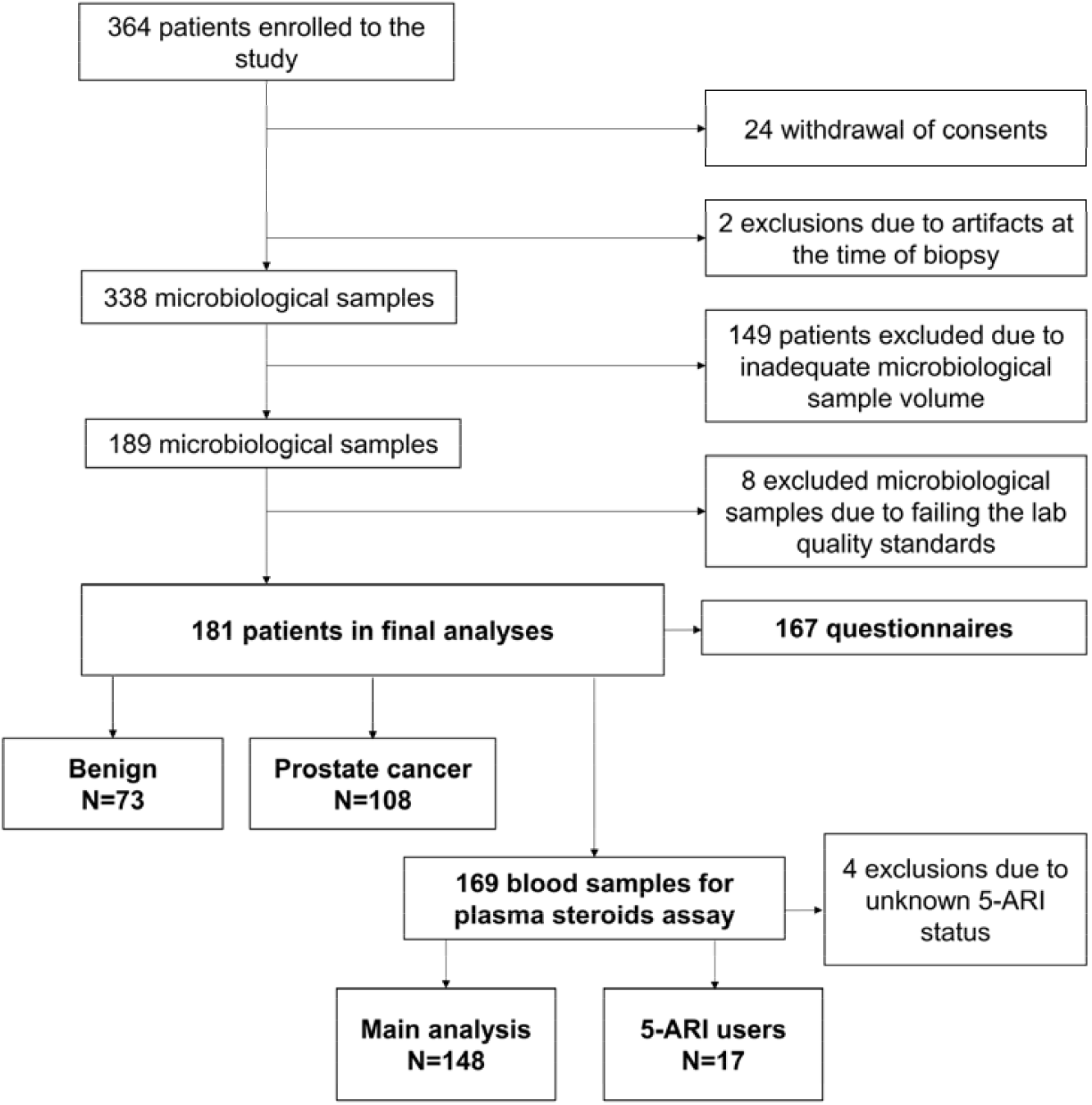
Study flowchart. 364 patients were enrolled in the study and after exclusions, 181 were included in the analyses. The available 169 blood samples were divided to main analysis group and 5-α-reductase inhibitor (5-ARI) users’ group.

The basic clinicopathological characteristics and the most significant confounding factors potentially affecting microbiota composition are presented in **Table 1**. Age, PSA, PSA-density, and prostate volume were significantly associated with cancer status and ISUP cancer grade. Potentially confounding factors were similar between groups except that the PCa group contained fewer active smokers (7%) than the benign group (13%). Smoking was not associated with cancer grade (Table 1).

**Table 1.** The clinicopathological characteristics of the subjects. Age, BMI, prostate cancer markers, 5-α-reductase inhibitor medication, recent antibiotic treatments, active smoking, special diet, recent travel, and inflammatory bowel diseases were analyzed.

| Characteristic | N | Benign |  | All cancers |  | All cancers vs Benign p-value | Cancer |  |  |  |  |  | Across the grade groups vs Benign p-value |
| --- | --- | --- | --- | --- | --- | --- | --- | --- | --- | --- | --- | --- | --- |
|  |  | Median | Q1-Q3 | Median | Q1-Q3 |  | ISUP Grade 1 |  | ISUP Grade 2-3 |  | ISUP Grade 4-5 |  |  |
|  |  |  |  |  |  |  | Median | Q1-Q3 | Median | Q1-Q3 | Median | Q1-Q3 |  |
| Age (years) | 167 | 62 | 55-67 | 68 | 62-72 | <0,001<sup>†</sup> | 63 | 58-68 | 68 | 65-73 | 69 | 66-72 | <0,001<sup>□</sup> |
| BMI (kg/m²) | 164 | 26,0 | 24,1-29,5 | 27,1 | 25,1-29,0 | 0,19<sup>†</sup> | 26,9 | 24,7-28,9 | 26,6 | 24,1-29,2 | 27,5 | 26,0-29,0 | 0,36* |
| PSA (µg/l) | 181 | 6,1 | 4,3-8,0 | 7,9 | 5,9-10,9 | <0,001<sup>†</sup> | 6,0 | 4,1-9,9 | 8,4 | 7,0-10,0 | 8,0 | 5,8-11,3 | <0,001* |
| PSA-density (ng/ml²) | 175 | 0,12 | 0,10-0,20 | 0,21 | 0,15-0,30 | <0,001<sup>†</sup> | 0,14 | 0,10-0,26 | 0,24 | 0,16-0,32 | 0,23 | 0,18-0,28 | <0,001* |
| free-PSA ratio (%) | 155 | 16 | 11-20 | 13 | 11-18 | 0,21<sup>†</sup> | 16 | 11-21 | 14 | 11-19 | 12 | 10-16 | 0,19* |
| Prostate volume (ml) | 175 | 44 | 33-62 | 36 | 29-48 | 0,003<sup>†</sup> | 39 | 31-49 | 35 | 28-52 | 36 | 31-43 | 0,031* |
|  |  | % |  | % |  |  | % |  | % |  | % |  |  |
| 5-α-reductase medication | 167 | 12 |  | 9 |  | 0,46<sup>‡</sup> | 8 |  | 4 |  | 18 |  | 0,18<sup>§</sup> |
| Antibiotic treatments within a year | 165 | 32 |  | 28 |  | 0,55<sup>‡</sup> | 30 |  | 20 |  | 39 |  | 0,30<sup>‡</sup> |
| Active smoking | 165 | 20 |  | 9 |  | 0,042<sup>‡</sup> | 13 |  | 9 |  | 7 |  | 0,20<sup>‡</sup> |
| Special diet | 165 | 2 |  | 3 |  | 1,000<sup>§</sup> | 8 |  | 2 |  | 0 |  | 0,26<sup>§</sup> |
| High risk traveling within a year¶ | 167 | 6 |  | 6 |  | 1,000<sup>§</sup> | 4 |  | 9 |  | 3 |  | 0,83<sup>§</sup> |
| Inflammatory bowel disease | 167 | 5 |  | 0 |  | 0,060<sup>§</sup> | 0 |  | 0 |  | 0 |  | 0,34<sup>§</sup> |

□ Kruskal wallis test
† Wilcoxon Rank sum test
‡ Pearson Chi square test
§ Fisher's exact test
□ finasteride/dutasteride
¶ Africa, Asia and South America

### α- AND β-DIVERSITY OF THE GUT MICROBIOTA

According to α-diversity indices Chao1 (p=0.7) and Shannon (p=0.3), no significant differences were found in bacterial community richness nor evenness (**Supplementary Figures S2-S4**). On the other hand, compositional dissimilarity differed between PCa cases in comparison to men without PCa (p=0.039, **Figure 2A**), but these differences were not reflected in the cancer grade (p=0.23, **Supplementary Figure S5**). The principal coordinate 1 (PCo 1) in Bray-Curtis analysis appears to consist of Prevotella 9 (10%), (**Figure 2B**), which was the most abundant single genus in the GM and associated with PCa in differential abundance analysis.

**Figure 2.**
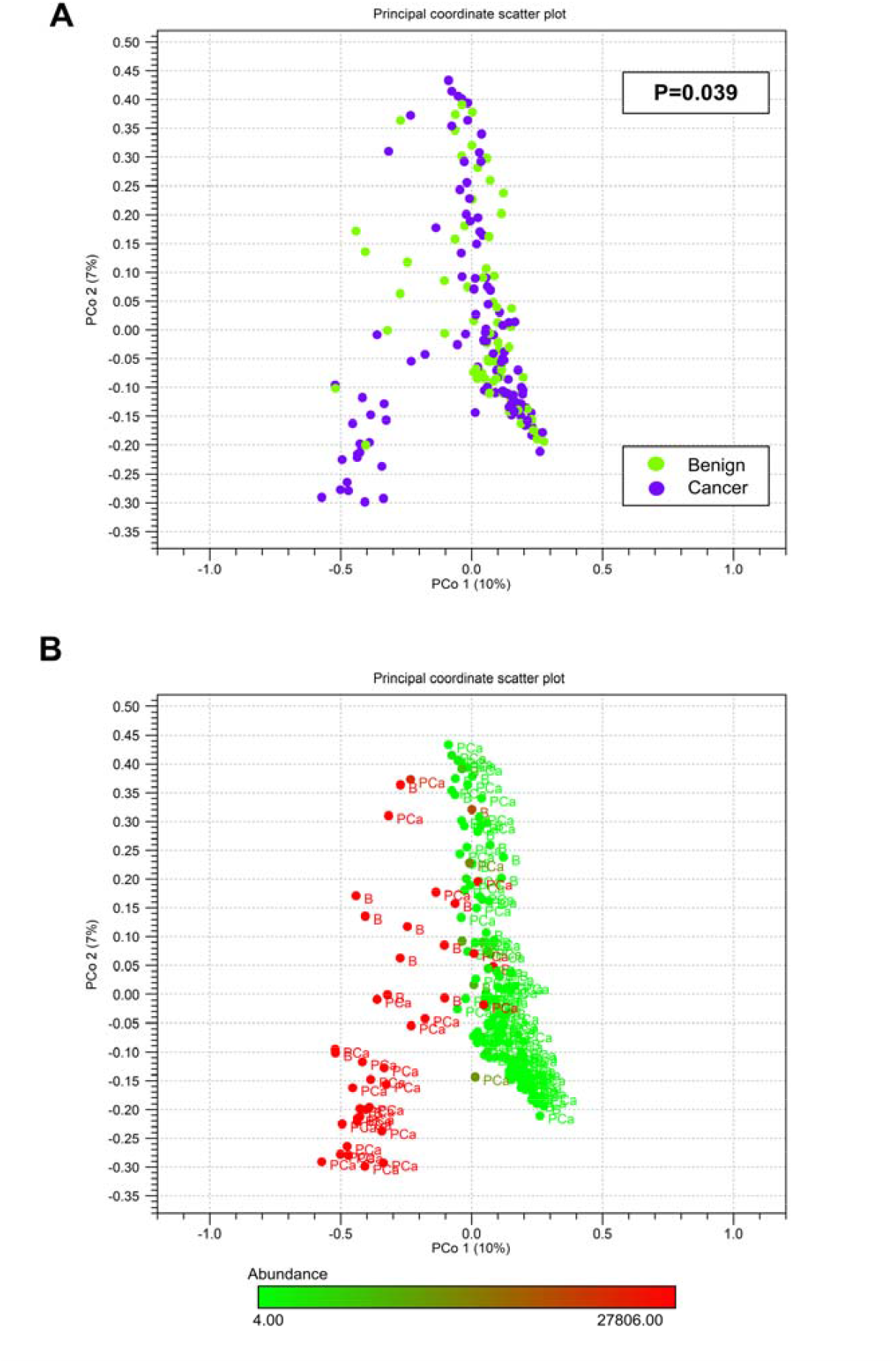
**A. Bray-Curtis dissimilarity of the gut microbiota between benign and prostate cancer (PCa) cases.** Microbiota of benign and cancer patients differs significantly in PERMANOVA analysis. **B. Prevotella 9 in the principal coordinate (PCo) scatter plot** 1 (10% of the calculated differences). Prevotella 9 abundance is also significantly elevated in PCa. Upper bound on the color scale is Q3 (27806) of cancer cases.

### OTU CLUSTERING AND DIFFERENTIAL ABUNDANCE ANALYSIS

The 181 successfully sequenced samples contained bacterial taxa from 21 different phyla. At the family level, OTU clustering showed *Prevotellaceae* becoming gradually more abundant with increasing cancer grade (**Figure 3**). Furthermore, differential abundance analysis showed that *<u>Alloprevotella, Prevotella 2</u>*, and *Prevotella 9* within the family *Prevotellaceae* were more abundant in the cancer group. Other significantly abundant genera belonged to families *Acidaminococcaceae, Christensenellaceae, Clostridiales vadinBB6*0, C*orynebacteriaceae, Enterobacteriaceae, Erysipelotrichaceae, Lachnospiraceae, Muribaculaceae, Ruminococcaceae, Synergistaceae* and Veillonellaceae. Genera higher and lower in PCa are presented in **Figure 4 (Supplementary Table S1)**. The microbial taxa, that had significantly different abundance in the cancer and benign subjects, had variable abundance correlation with the different PCa ISUP grades. E.g., some microbes were especially abundant in low grade cancers (e.g. *Acidaminococcus*), while others were higher only in men with higher grade cancers (e.g. UBA1819, [*Clostridium*] *innocuum*) **(Supplementary Figures S6A-D)**.

**Figure 3.**
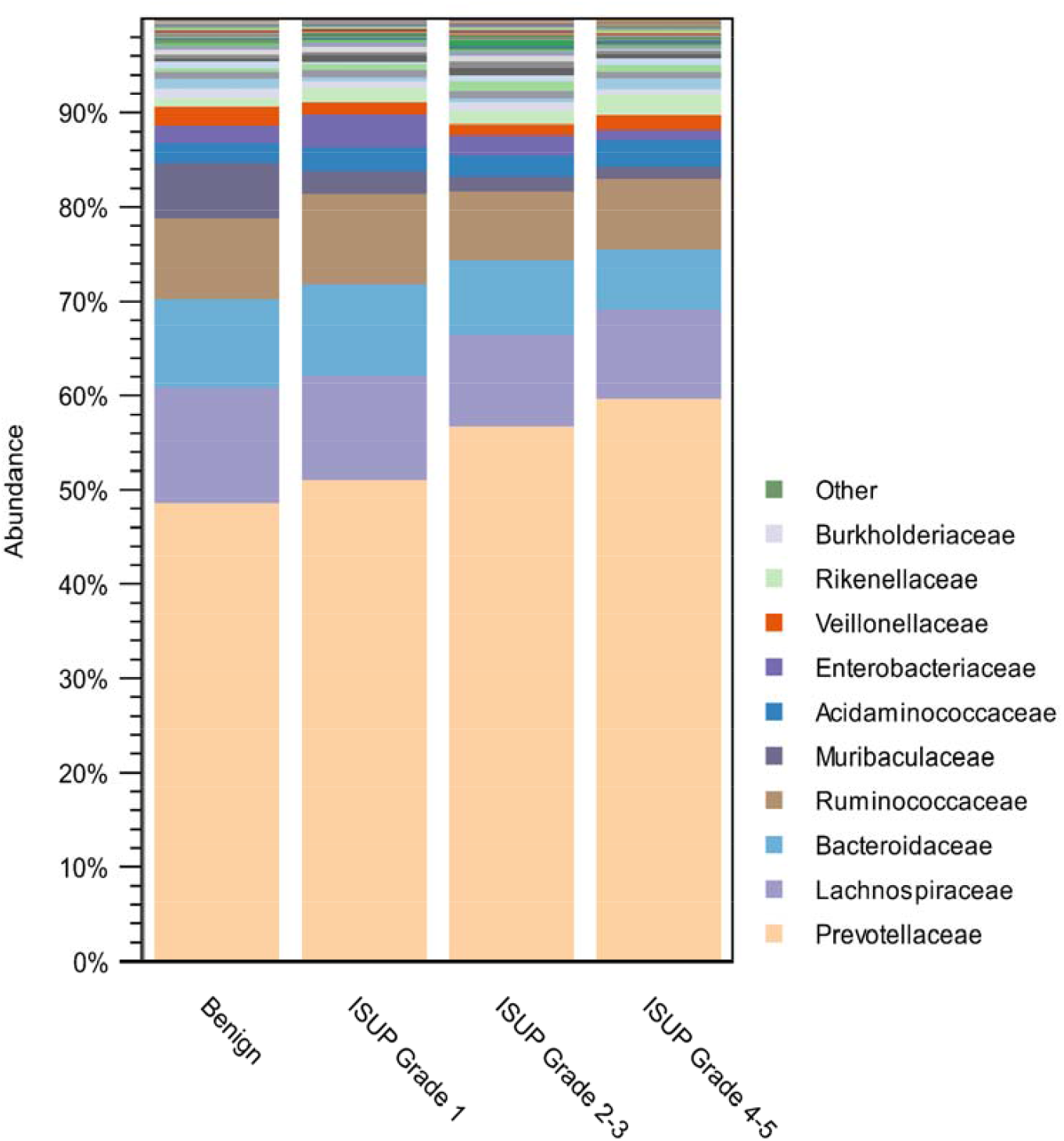
OTU table of the core microbiome at the family level. Family Prevotellaceae abundance elevates with cancer severity.

**Figure 4.**
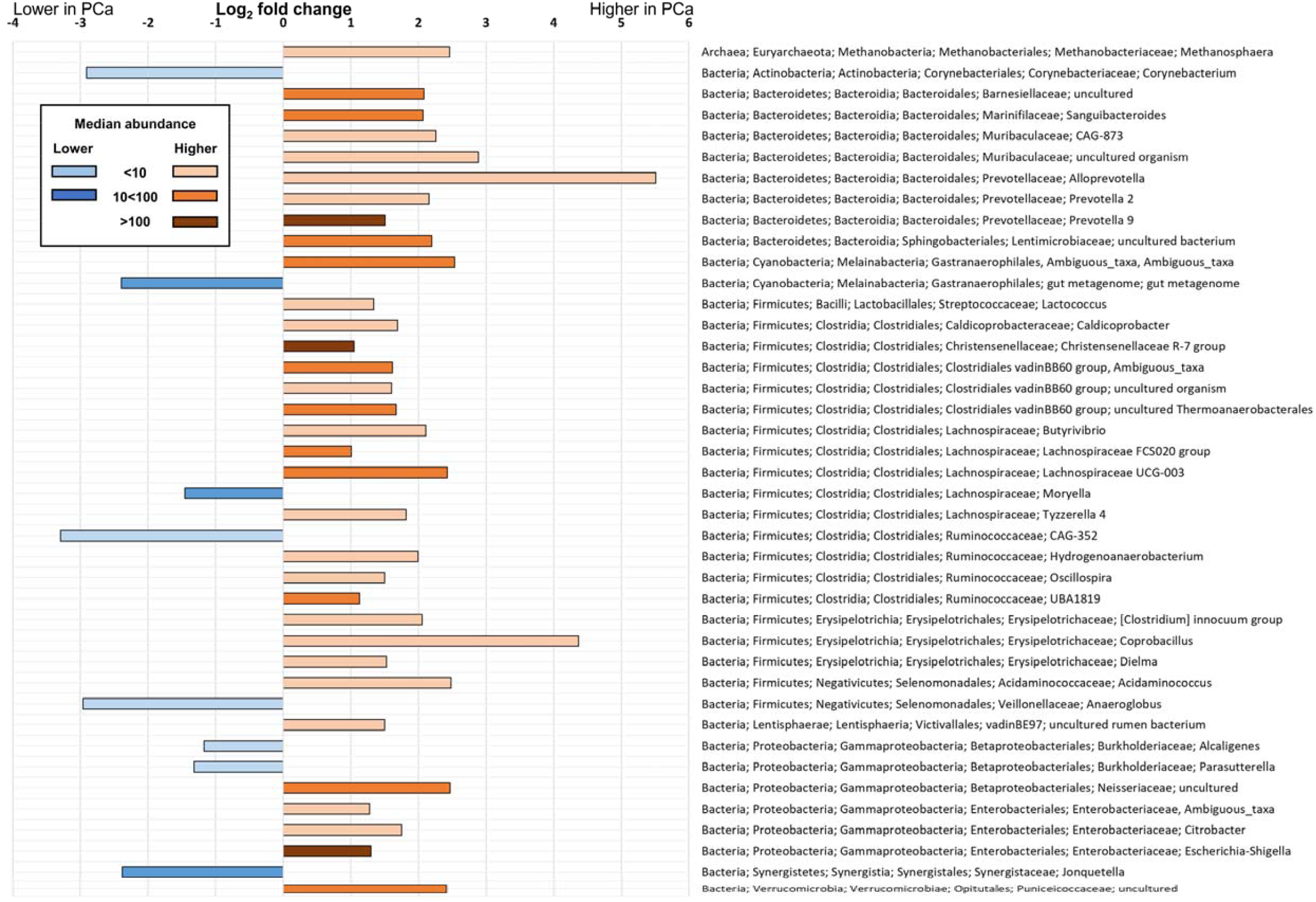
Log_2_ fold changes of microbial genera associated with PCa. Genera higher in prostate cancer cases (peach-orange-brown shades) includes known pathogens as *Escherichia-Shigella* and genera that have been formerly associated with prostate cancer, such as *Erysipelotrichaceae* members [*Clostridium*] *innocuum, Coprobacillus* and *Dielma*. Genera lower in the cancer cases (blue shades) include CAG-352, *Anaeroglobus* and *Corynebacterium*. The shade of the color indicates the dominance of the genus by median abundance in the whole cohort. Genera are presented in the taxonomical order.

### PREDICTIVE ANALYSIS OF BACTERIAL FUNCTIONAL GENES (PICRUSt)

Three KEGG Pathways were significantly different between the cancer and benign groups; mineral absorption (copper chaperone, p=0.008), steroid hormone biosynthesis (5-α-reductase, 5-AR, p=0.022), and retinol metabolism (p=0.042) had significantly higher values in the PCa group compared to men without cancer (**Table 2, Supplementary Table S2**).

**Table 2.** Predicted bacterial gene products associated with PCa.

| KEGG Pathway | KEGG Orthologs | Benign |  | Cancer |  | p-value |
| --- | --- | --- | --- | --- | --- | --- |
|  |  | Median | 95% CI<br>[LB; UB] | Median | 95% CI<br>[LB; UB] |  |
| Mineral absorption | Copper chaperone (95%) | 2522 | [1633; 3716] | 4044 | [2325; 6090] | **0.008** |
|  | Heme oxygenase 1 (5%) |  |  |  |  |  |
| Steroid hormone biosynthesis | 5-α-reductase 1 (45%) | 8457 | [4514; 10952] | 11717 | [8508; 14660] | **0.022** |
| | 5-α-reductase 2 (54%) | | | | | |
| | 3-α-hydroxysteroid hydrogenase (1%) | | | | | |
| Retinol metabolism | Alcohol dehydrogenase (67%) | 21605 | [18680; 23784] | 23174 | [21171; 26117] | **0.042** |
|  | Alcohol dehydrogenase propanol preferring (5%) |  |  |  |  |  |
|  | All-trans-retinol 13,14-reductase (20%) |  |  |  |  |  |
|  | S-(hydroxymethyl) glutathione dehydrogenase/alcohol dehydrogenase (8%) |  |  |  |  |  |

### PLASMA STEROIDS

The possible systemic effects of steroid hormone metabolism suggested by the KEGG pathways (PICRUSt) were investigated. Firstly, among 5-ARI users (n=17) the plasma DHT (p<0.0001) and DHT/T ratio (p=0.0002) were significantly lower compared to non-users, as expected (**Figure 5**). Secondly, within this subgroup of 5-ARI users, the higher predicted microbial 5-AR was associated with lower estrone (p=0.027) and estradiol (p=0.059) levels (**Figure 6**). Among non-5-ARI users (N=148), plasma testosterone levels negatively associated with predicted microbial 5-AR (Spearman correlation −0.138, p=0.095, Wilcoxon rank sum p=0.030) (**Figure 7**). In other words, plasma testosterone levels were lower among subjects with predicted elevated GM 5-AR levels when compared to men with predicted low GM 5-AR levels.

**Figure 5.**
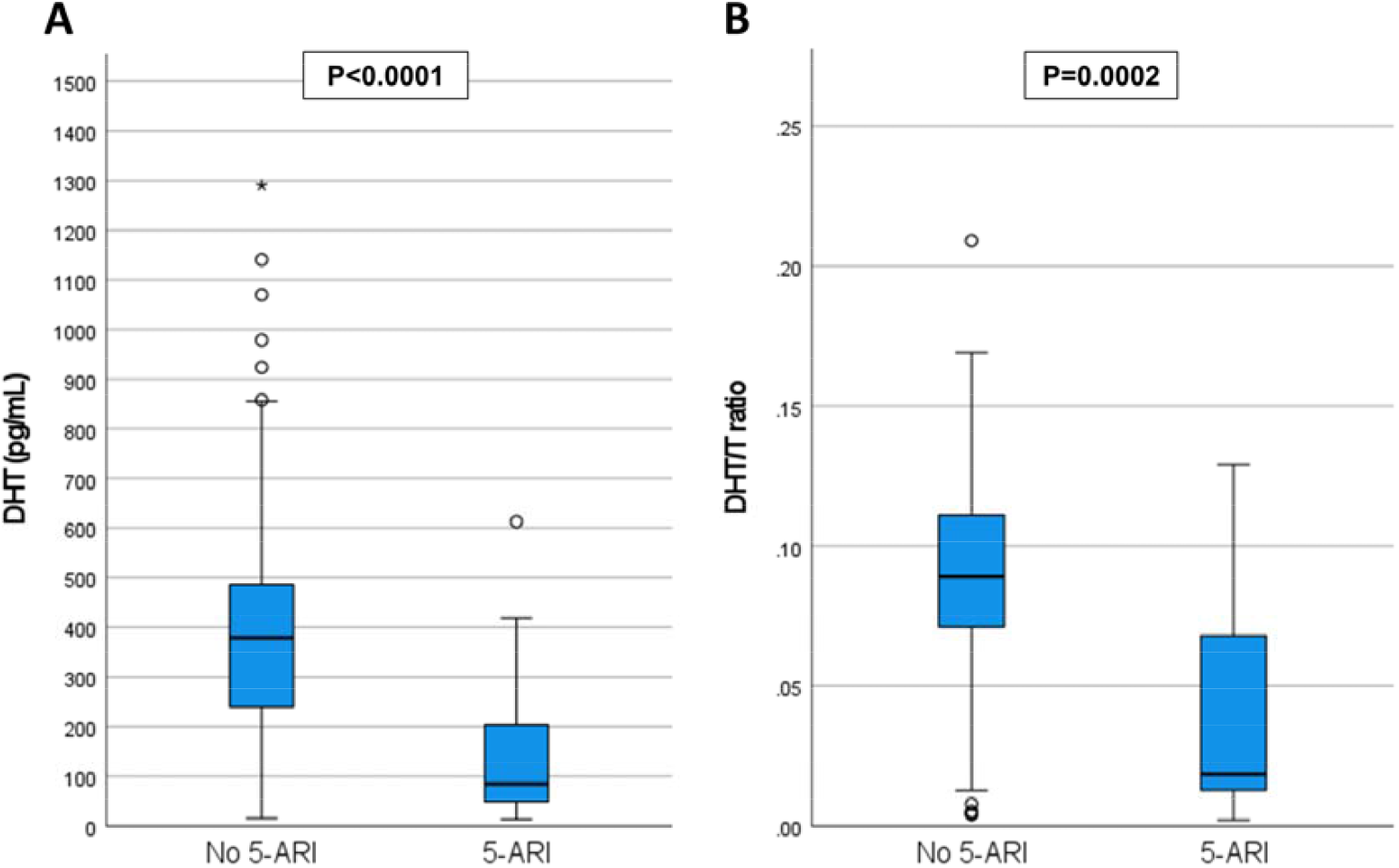
**A. DHT concentration in plasma by 5-α-reductase inhibitor (5-ARI) medication status.** Among 5-ARI users, DHT levels are significantly lower in plasma*. **B. DHT/T ratio in plasma by 5-ARI medication status**. Among 5-ARI users, DHT/T ratio is significantly lower in plasma*. (*Wilcoxon Rank Sum test)

**Figure 6.**
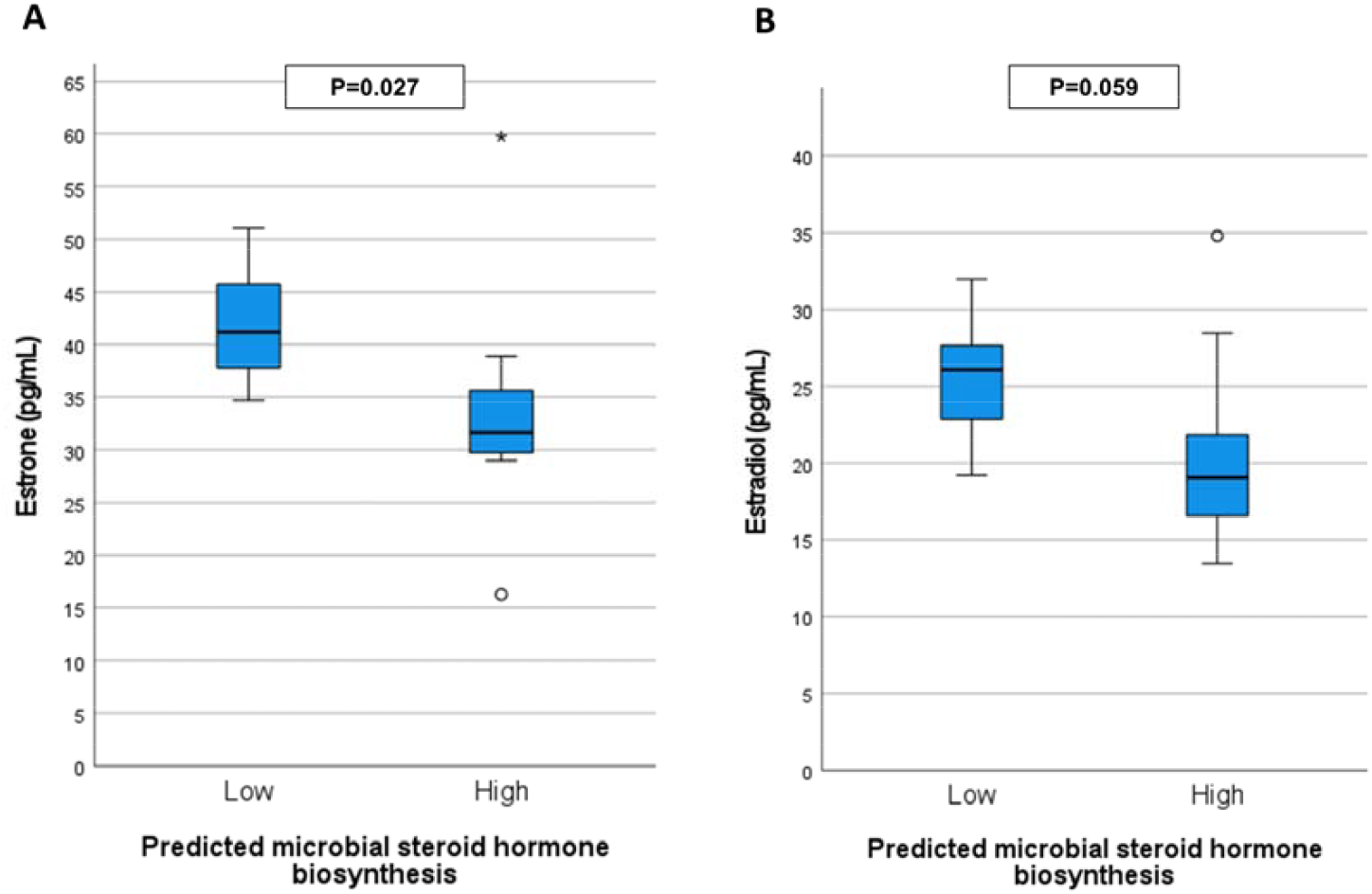
**A. Estrone concentration in plasma by Predicted microbial steroid hormone biosynthesis among 5-α-reductase inhibitor (5-ARI) users.** Plasma estrone concentration is significantly lower in high (above median) microbial steroid hormone biosynthesis group*. **B. Estradiol concentration in plasma by Predicted microbial steroid hormone biosynthesis among 5-ARI users**. Plasma estradiol is lower in high (above median) microbial steroid hormone biosynthesis group*. (*Wilcoxon Rank Sum test)

**Figure 7.**
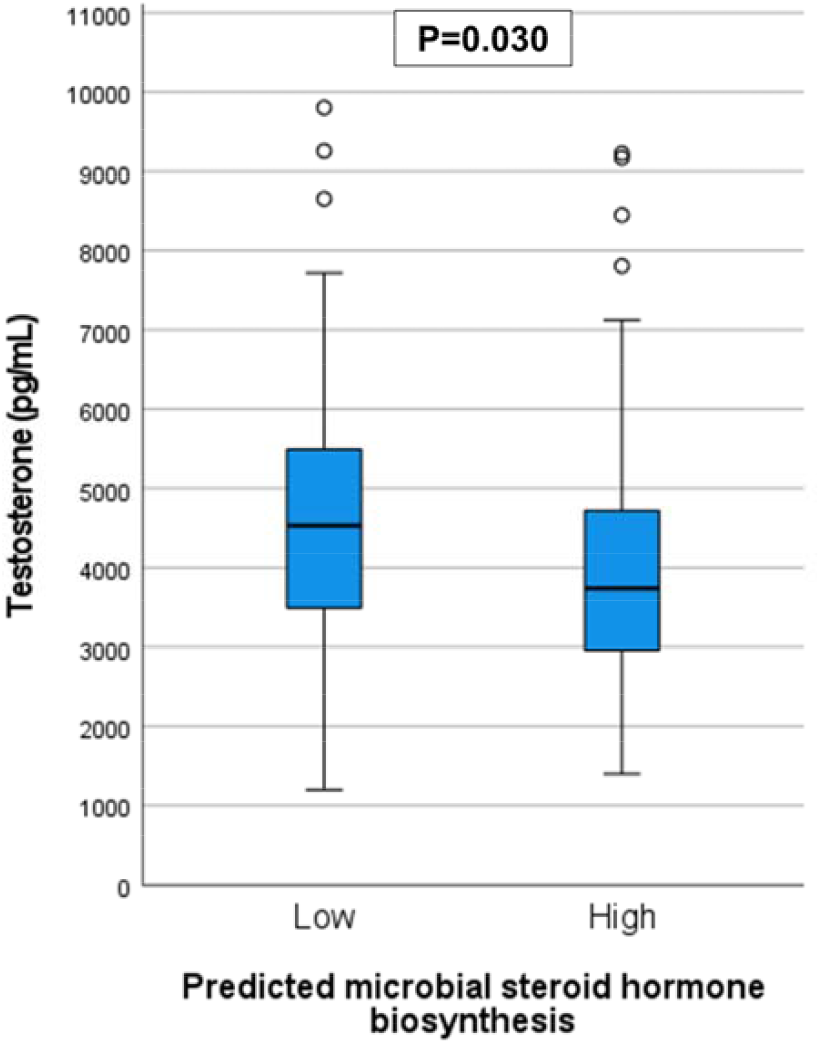
Testosterone concentration in plasma by Predicted microbial steroid hormone biosynthesis. Plasma testosterone levels are significantly lower in patients that have high (above median) microbial steroid hormone biosynthesis without adjusting for the cancer status. (Wilcoxon Rank Sum test)

## DISCUSSION

This study evaluated a potential link between GM and PCa using GM profiles of 181 patients with suspected PCa. The 108 patients in whom PCa was diagnosed had significant differences in their GM signatures compared to the remaining 73 patients. Predictive functional analyses suggest that differences in the steroid hormone, copper, and retinol pathways could be the metabolic consequences of the altered microbiota. The hormone dependency of PCa could render it susceptible to alterations in steroid hormones caused by the GM, which were also studied. Men having a predicted alteration in gut steroid hormone metabolism, measured as a predicted higher microbial 5-AR, also had patterns of plasma steroid concentration that differed from those men not having this predicted elevated level.

In the detailed analyses, the abundance of family *Prevotellaceae* within phylum Bacteroidetes got gradually higher among cancer cases with cancer severity according to ISUP grade. Members of *Prevotellaceae* family are known to belong to a healthy core microbiota within the gut and have been shown to be affected especially by diet since they are able to degrade complex plant polysaccharides.^20^ However, *Prevotella* spp. have recently been shown to be enriched in both in colorectal tumours and mucosal microbiota of CRC patients, thus implying a possible role of these species in other cancers.^21,22^ Interestingly, *Prevotella 9* was the most abundant bacterial genus detected in the whole study cohort, but to date the biological significance of *Prevotella 9* in the context of microbiota and host health has not been elucidated.

Furthermore, differential abundance analysis revealed other interesting bacteria enriched in the cancer group, including genera from core microbiota to less abundant ones. A core member, *Escherichia-Shigella* from *Enterobacteriaceae* family, includes the well-known uropathogen *E. coli* that has colibactin mediated genotoxic properties and has been reported in PCa patients previously.^7,23^ In our study, *Clostridiales* were widely represented higher in PCa cases, which have been proposed earlier.^9^ On the other hand, *Erysipelotrichaceae* family has been suggested to play a role in the pathogenesis of several diseases, such as IBD and metabolic disorders, even though it is usually a minor member of the GM community, as in this study.^24^ To conclude, several microbes higher in PCa have been previously linked to another diseases, highlighting the disease promoting properties of the GM.

It is of great interest that the PICRUSt analysis indicated elevated steroid hormone biosynthesis pathways in the GM of the cancer cases, since it is well known that 5-AR reduces testosterone to dihydrotestosterone (DHT) feeding prostate cancer. Formerly, higher steroid hormone biosynthesis in microbiota of patients with androgen deprivation therapy (n=9) when compared to men without therapy (n=16) has been discovered.^25^ Furthermore, the contribution of the GM in the rate of PCa tumor growth, and progression of castration resistance through steroid hormones in animal models as well as in human samples was shown recently.^26^ These studies suggest that steroid metabolism of GM is associated with therapy response. However, our study suggests that microbiota enriched 5-AR activity may have more profound effects in prostate carcinogenesis. In addition, we demonstrated that the systemic steroid hormone levels associated with altered predicted microbial 5-AR activity in a biologically rational manner, since higher 5-AR lowers testosterone. The potential mechanistic explanation for altered steroid hormone metabolism caused by GM clearly warrants further studies.

The other potential metabolic pathways noted in our study were copper and retinol metabolism that are less studied in PCa. Copper chaperone is a protein that ferries copper to cellular organelles and interestingly, accumulation of copper has been reported in tumor cells, including PCa cells.^27^ Androgen receptor activation enhances copper uptake, again suggesting that hormonal pathways are involved in altered GM metabolism.^28^ In line with our findings, higher retinol (vitamin-A derivate) concentrations have been associated with elevated PCa risk and genetic variants in retinol pathways have been associated with PCa.^29,30^

The main limitation of our study is the lack of sample size estimations and power calculations, but this would have been impossible to carry out without prior studies. Furthermore, low stool content in the sampling swabs, resulting in unsatisfactory amount of bacterial DNA for NGS in 149 of 338 samples, could have been avoided by a better fecal sampling method. Furthermore, our study did not include a microbial follow-up which would be needed to investigate the actual progression of microbiota and prostate cancer. The PICRUSt analysis was performed with Greengenes database, which may differ from the results acquired with SILVA. One should also note that the study was limited to subjects with relatively low risk cancer suspicion, which may potentially dilute the results as cases with advanced or metastatic tumors were not included. The strengths of our study include the prospective design, detailed prospective data collection and that, to our knowledge, this is the largest reported study on the subject.

## CONCLUSIONS

In this study we discovered previously unreported differences in GM components between prostate cancer patients and benign subjects. In a predictive analysis microbial steroid hormone biosynthesis, mineral absorption, and retinol metabolism were potential carcinogenic pathways. Moreover, elevated predicted microbial 5-AR associated with lower testosterone in plasma. These findings could explain the observed association of lifestyle, geography and PCa incidence.

## Data Availability

Clinical data available at http://petiv.utu.fi/multiimprod

## ACKNOWLEDGEMENTS

This work was supported by grants from The Cancer Foundation Finland and TYKS-SAPA research fund of Turku University Hospital.

We thank Peter B. Dean, MD D. Med. Sci. (University of Turku, Turku, Finland) for helping with the manuscript revision.

## ABBREVIATIONS

PCa: Prostate cancer
GM: Gut microbiota
ISUP: International Society of Urological Pathology
BMI: Body mass index
IBD: Inflammatory bowel disease
OTU: Operational Taxonomical Unit
PICRUSt: Phylogenetic Investigation of Communities by Reconstruction of Unobserved States
DHT: Dihydrotestosterone
DHEA: Dehydroepiandrosterone
T: Testosterone
17-α-OHP: 17-α-hydroxyprogesterone 5-AR 5-α-reductase
5-ARI: 5-α-reductase inhibitor

## DATA SHARING STATEMENT

All de-identified MRI data and the study protocol is available on the Multi-IMPROD site (http://petiv.utu.fi/multiimprod). All microbiological data will be available for bona fide researchers who request it from the authors.

