## Supplementary material for "Gut microbiota affects prostate cancer risk through steroid hormone biosynthesis"

This appendix has been provided by the authors to give readers additional information about their work.

#### LIST OF INVESTIGATORS

Sofia Kalinen[1,2],B.M., Teemu Kallonen[2,3],Ph.D., Marianne Gunell[2,3],Ph.D., Otto Ettala[4], M.D., Ph.D., Ivan Jambor[5], M.D., Ph.D., Juha Knaapila[4], M.D., Ph.D., Kari T. Syvänen[4], M.D., Ph.D., Pekka Taimen[6,7], M.D., Ph.D., Matti Poutanen[6,8,9], Ph.D., Claes Ohlsson[9], M.D., Ph.D., Hannu J. Aronen[5], M.D., Ph.D., Helena Ollila[10], M.Sc, Sami Pietilä[11],M. Sc., Laura L. Elo[6,11], Ph.D., Tarja Lamminen[4], Ph.D., Antti J. Hakanen[2,3], M.D., Ph.D., Eveliina Munukka[3,12], Ph.D., Peter J. Boström[4], M.D., Ph.D., and the Multi-IMPROD Study group*

[1] Research Center for Infections and Immunity, Institute of Biomedicine, University of Turku, Turku, Finland
[2]Department of Clinical Microbiology, Turku University Hospital, Turku, Finland
[3]Microbiome Biobank, University of Turku, Turku, Finland
[4]Department of Urology, Turku University Hospital and University of Turku, Turku, Finland
[5]Department of Radiology, Turku University Hospital and University of Turku, Turku, Finland
[6]Institute of Biomedicine, University of Turku, Turku, Finland
[7]Department of Pathology, Turku University Hospital, Turku, Finland
[8]Centre for Integrative Physiology and Pharmacology, University of Turku, Turku, Finland
[9]Department of Internal Medicine and Clinical Nutrition, Institute of Medicine, Sahlgrenska Academy, University of Gothenburg, Gothenburg, Sweden
[10]Turku Clinical Research Centre, Turku University Hospital, Turku, Finland
[11]Turku Bioscience Centre, University of Turku and Åbo Akademi University, Turku, Finland
[12]Biocodex: Biocodex Nordics, Espoo, Finland

*__Multi-IMPROD study group:__

From the Multi-IMPROD study group, Included in author list: Ivan Jambor, Otto Ettala, Juha Knaapila, Pekka Taimen, Kari T. Syvänen, Tarja Lamminen, Hannu Aronen, Peter J. Boström

The study group also includes following investigators:
Janne Verho, Department of Radiology, Turku University Hospital and University of Turku, Turku, Finland
Aida Steiner, Department of Radiology, Turku University Hospital and University of Turku, Turku, Finland
Esa Kähkönen, Department of Urology, Turku University Hospital and University of Turku, Turku, Finland
Ileana Montoya Perez, Department of Computing, University of Turku, Turku, Finland
Marjo Seppänen, Department of Surgery, Satakunta Central Hospital, Pori, Finland
Antti Rannikko, Department of Urology, Helsinki University, and Helsinki University Hospital, Helsinki Turku, Finland
Outi Oksanen, Department of Radiology, Helsinki University Hospital, Helsinki, Finland
Jarno Riikonen, Department of Urology, Tampere University Hospital, Tampere, Finland
Sanna-Mari Vimpeli, Department of Radiology, Tampere University Hospital, Tampere, Finland
Harri Merisaari, Department of Radiology, University of Turku
Markku Kallajoki, Department of Pathology, Turku University Hospital
Tuomas Mirtti, Department of Pathology, University of Helsinki, Helsinki, Finland
Jani Saunavaara, Department of Radiology, University of Turku, Turku Finland

### SUPPLEMENTARY RESULTS

#### Sample quality and sequencing batch effect

β-diversity measures and principal coordinate plots can be used to assess quality. In this study all used metrics showed eight diverging samples in principal coordinate plot (**Figure S1A-C**). Those eight samples had been extracted in the same batch.

Sequencing batch effect is seen on the PCo1 (18%) (**Figure S1C**). The batch effect was not seen on Weighted Uni Frac nor Bray-Curtis. (**Figure S1A-B**) Unweighted Uni Frac describes the absence/presence of microbes in the sample whereas weighted Uni Frac and Bray Curtis dissimilarity also take the abundance into account. According to these results the use of Weighted Uni-Frac and Bray-Curtis is acceptable.

#### Rarefaction curves

Number of reads and the Chao1 and Shannon entropies of the samples. The rarefaction level 10527 was set for the analyses as all the samples could meet that level (**Figure S2**).

#### α-Diversity

Shannon index and Chao1 metric did not show significant differences between the groups (**Figure S3-4**).

#### β-Diversity

Bray-Curtis dissimilarity between ISUP Grade Groups in principal coordinate plot. Statistical significance was tested with PERMANOVA with 99999 permutations. There were no significant differences between ISUP grade groups (**Figure S5).**

#### Differential abundance analysis

Table of significantly differentially abundant genera between cancer and benign cases with taxonomy information is presented in **Table S1.**

This was not a follow up study. However, differential abundance analysis was performed with ISUP grade groups to find out the microbiota associated with the prostate cancer severity. Several bacterial genera trend in the similar manner across the grade groups. The bacteria genera could be divided to initial, lower abundance in cancer and higher abundance in cancer according to $\log_2$ fold changes compared to benign. *Escherichia-Shigella* and *Lactococcus* were mostly higher in ISUP grade group 2-3. (**Figure S6A-D**). Paired $\text{Log}_2$ fold changes between the cancer grade versus benign were calculated for the significant (FDR-corrected) genera in ANOVA-like test.

#### PICRUSt

PICRUSt KEGG Pathways within P<0.10 are in the **Table S2**.

#### SUPPLEMENTARY FIGURES

**Figure S1**

**A**

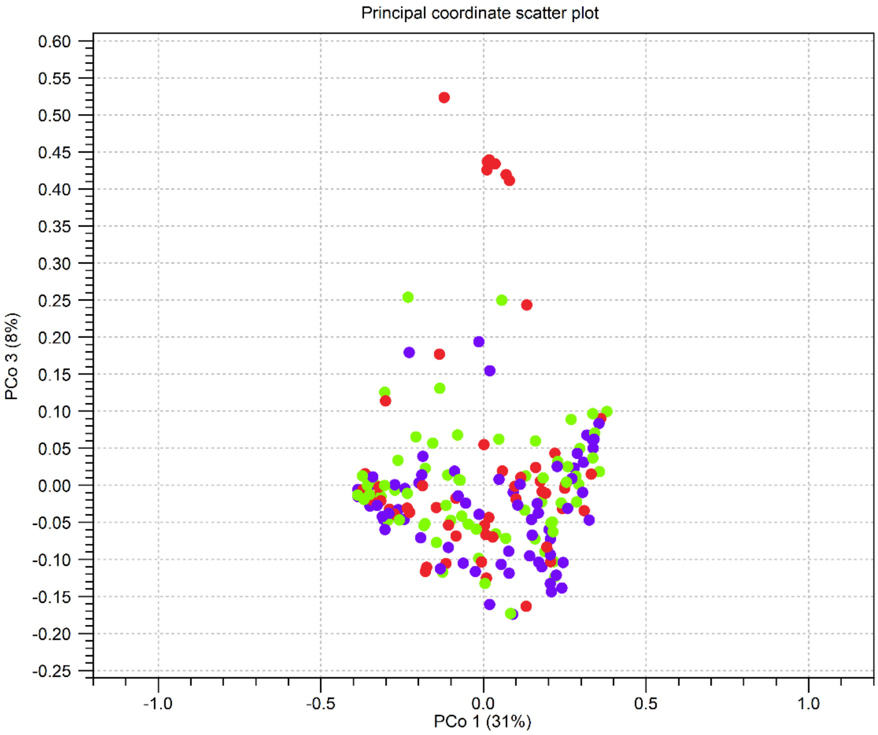

**B**

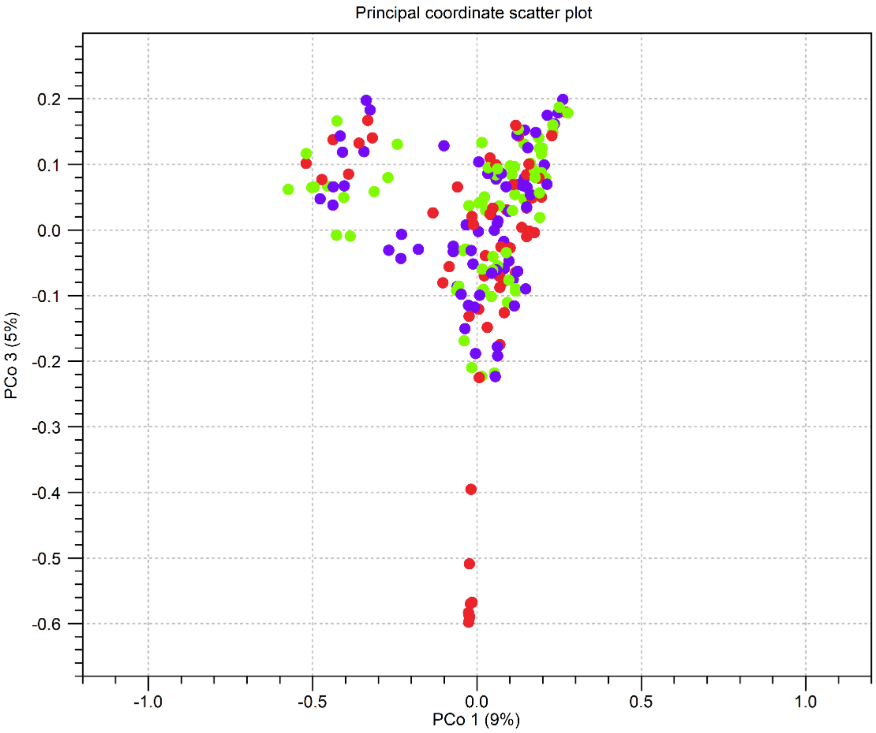

C

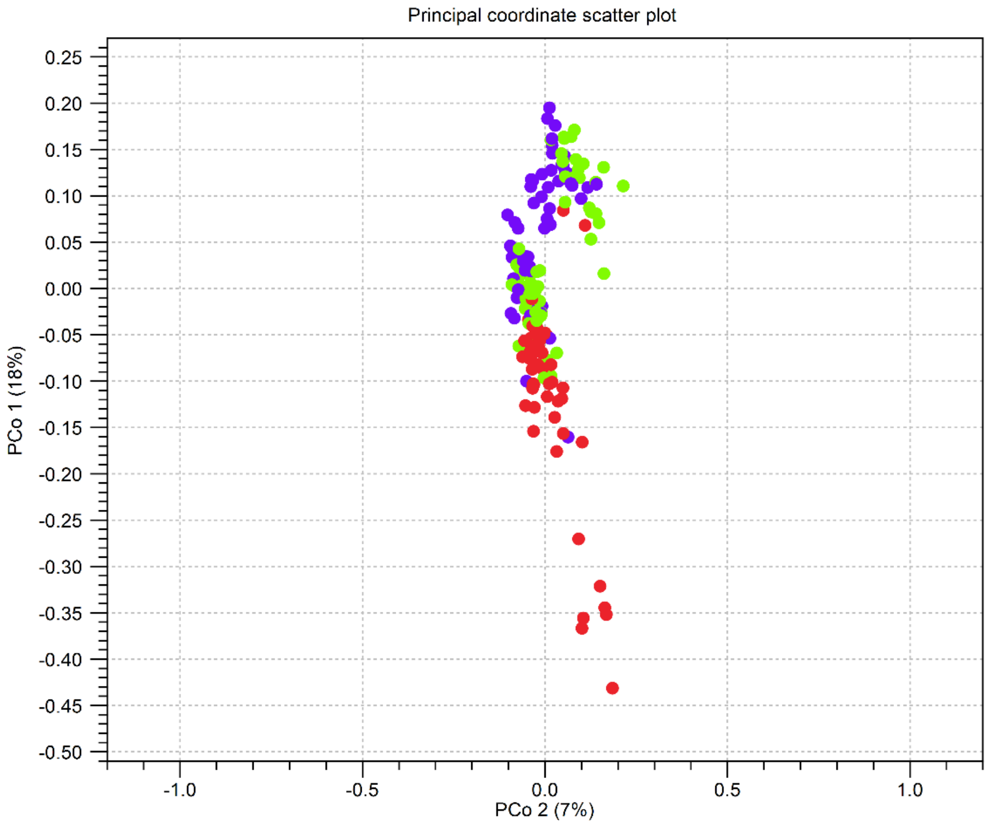

**Figure S1 Sample quality and sequencing batch in principal coordinate plots. A) Weighted Uni Frac B) Bray-Curtis. C) Unweighted Uni Frac. Samples were sequenced in three batches (red, green, purple). 8 samples (red) did not meet the quality standards. These samples were in the same DNA extraction batch and were removed from the further analyses.**

**Figure S2**

A

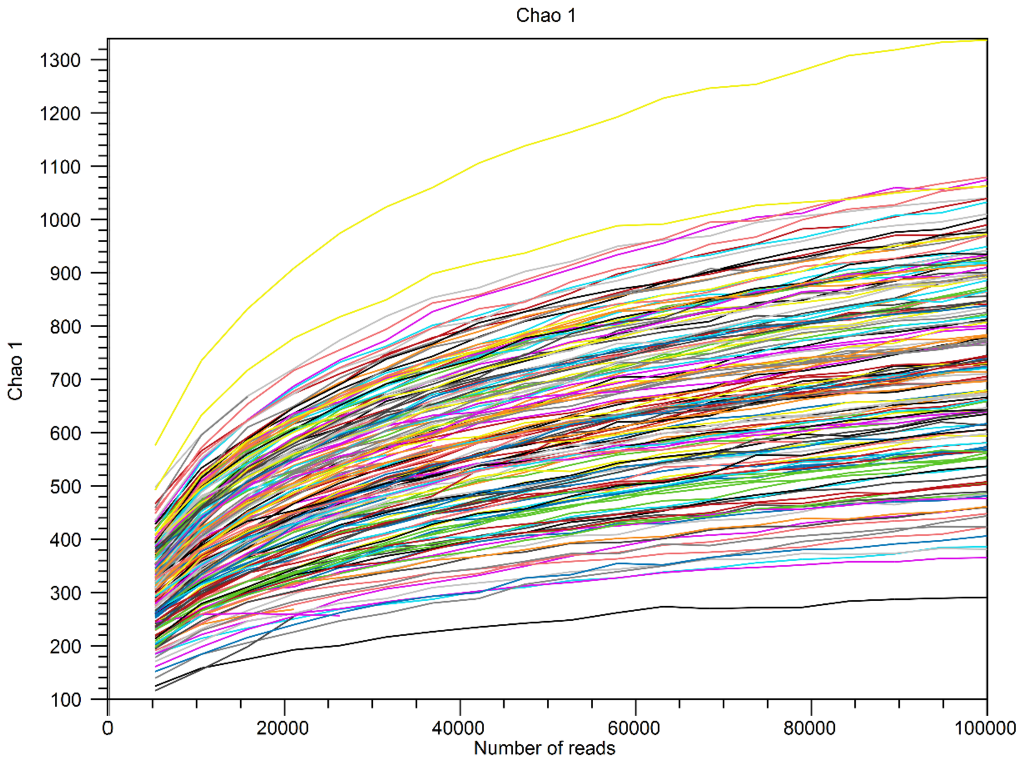

B

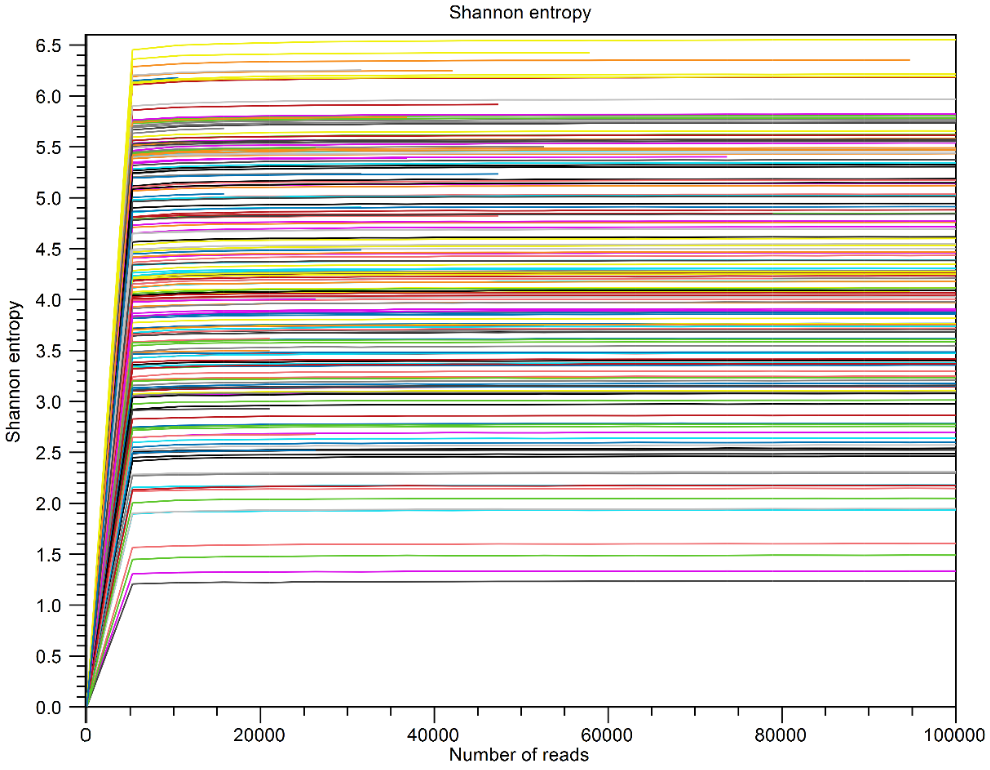

**Figure S2 Number of reads and α-diversity indices A: Chao1 B: Shannon entropy.**

**Figure S3**

**A**

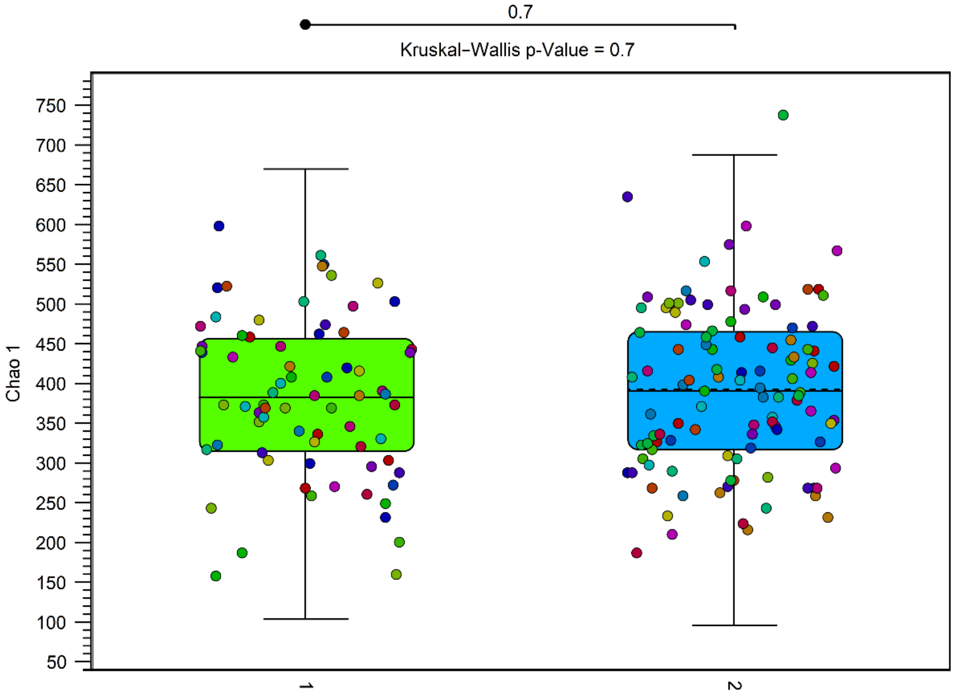

**B**

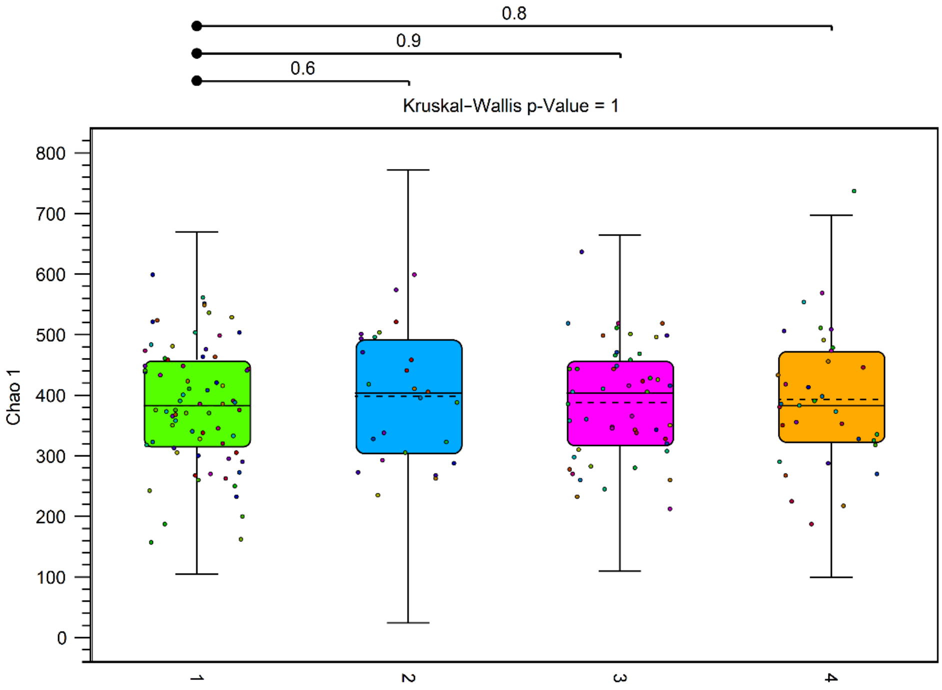

**Figure S3 Chao1 between groups. There were no significant differences between the groups. A: 1= Benign, 2=Cancer. B: 1=Benign, 2=ISUP 1, 3=ISUP 2-3, 4=ISUP 4-5.**

**Figure S4**

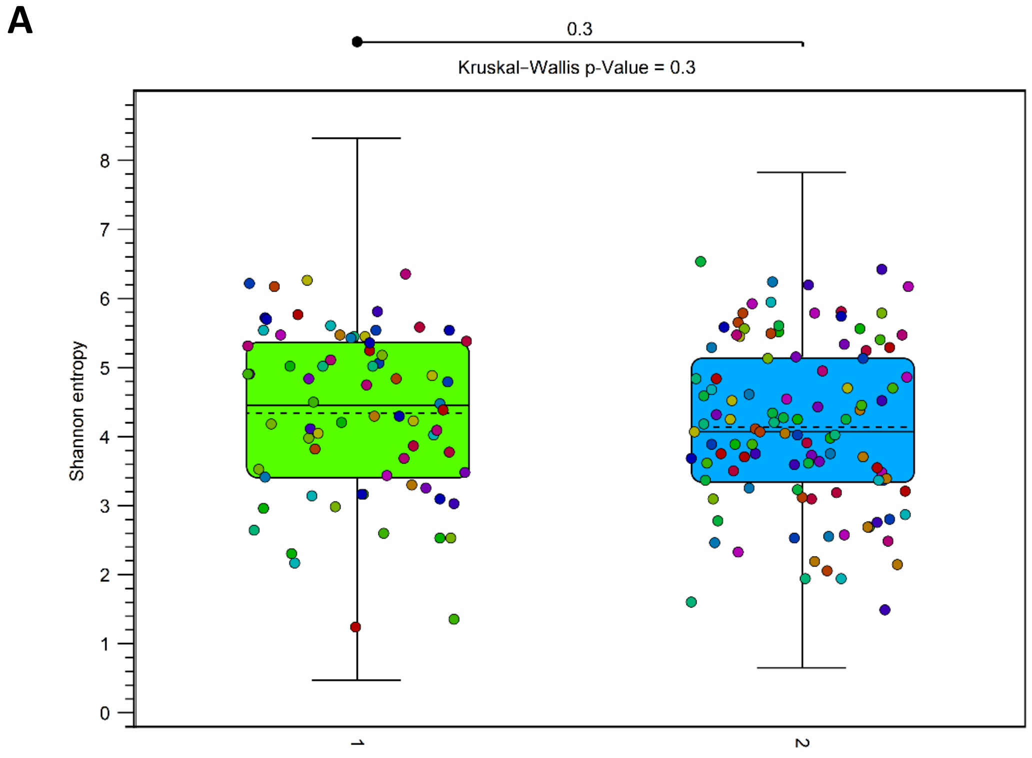

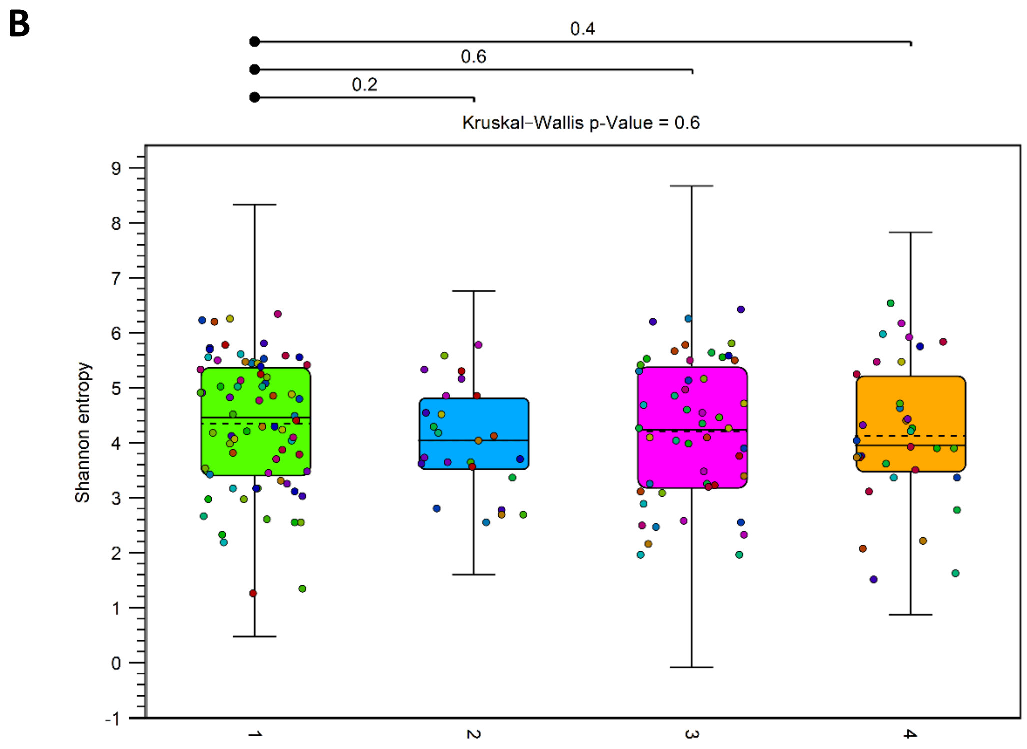

**Figure S4 Shannon entropy between groups. There were no significant differences between the groups. A: 1=Benign, 2= Cancer, B: 1=Benign, 2=ISUP 1, 3=ISUP 2-3, 4=ISUP 4-5.**

**Figure S5**

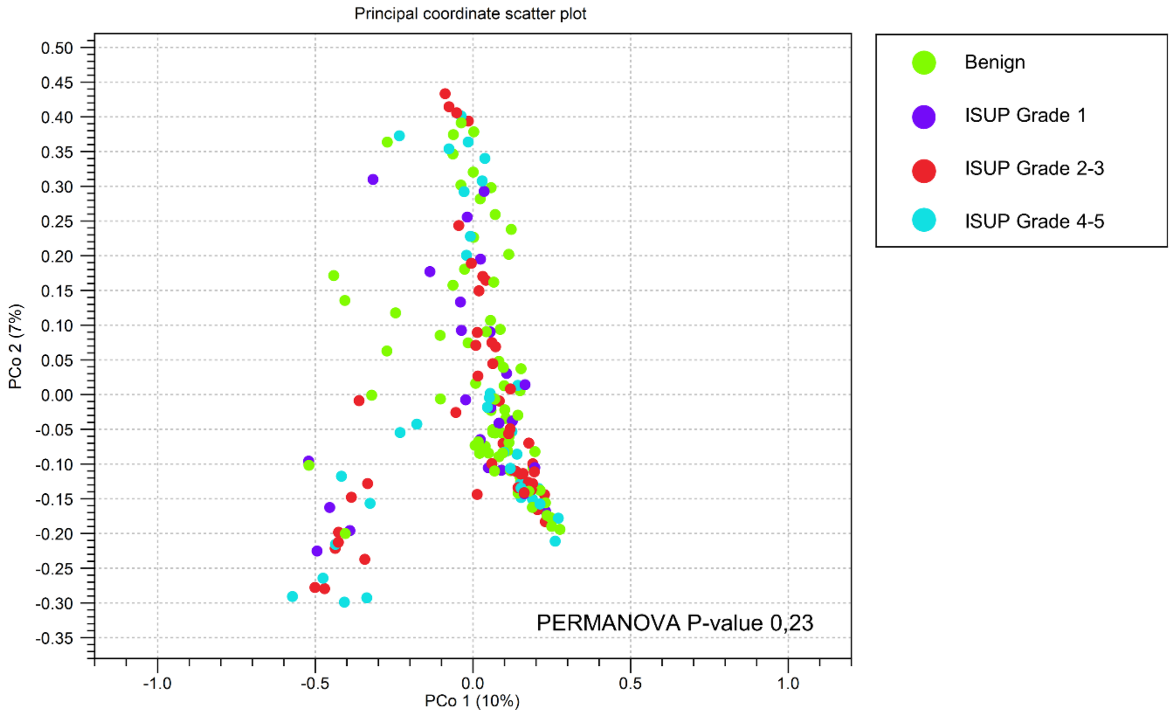

**Figure S5 β-Diversity of ISUP Grade Groups in principal coordinate plot. There were no significant differences between the ISUP grade groups.**

**Figure S6**

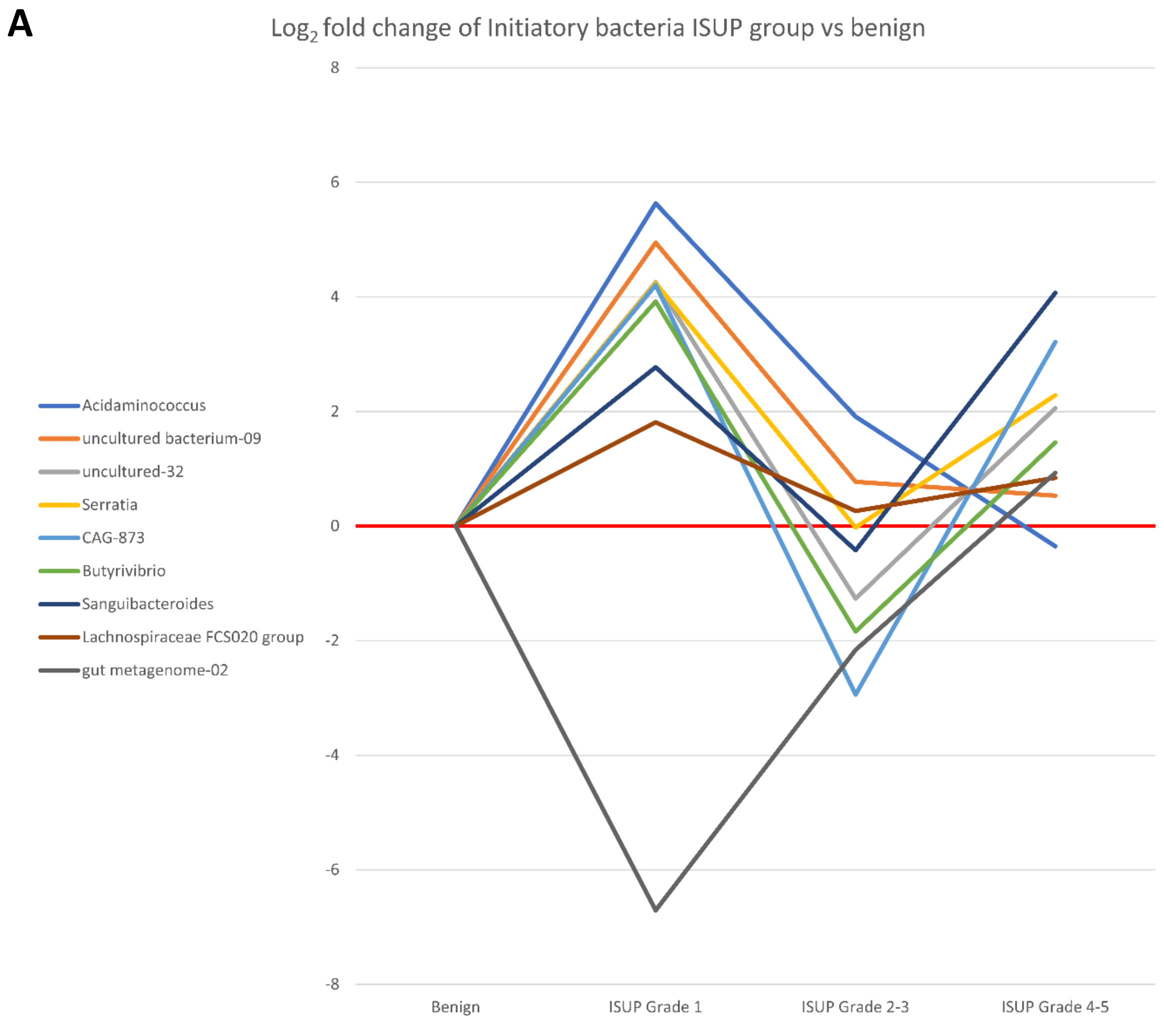

**Figure S6A $Log_2$ fold changes of potentially cancer initiating bacteria according to ISUP grade groups.**

**B**

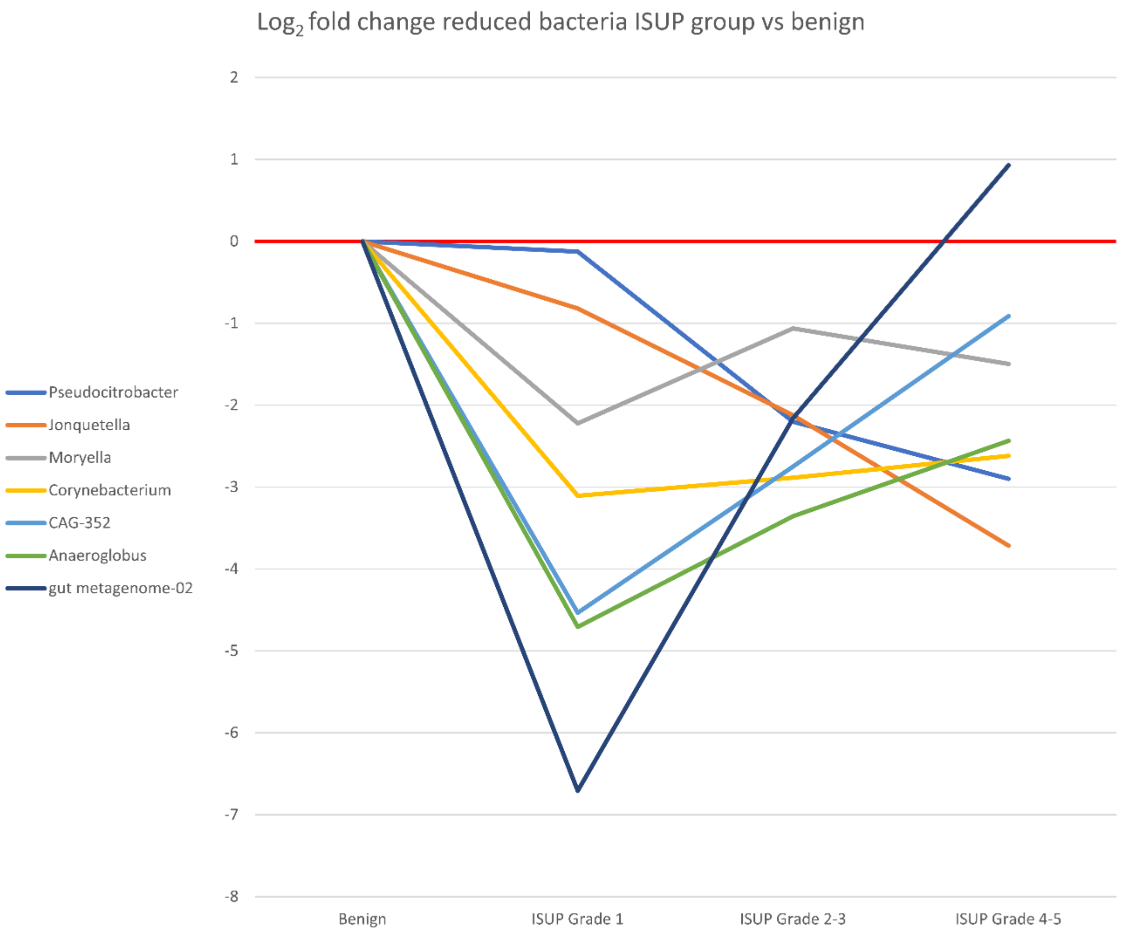

**Figure S6B Log$_2$ fold changes of lower abundance bacteria in prostate cancer according to ISUP grade groups.**

**C**

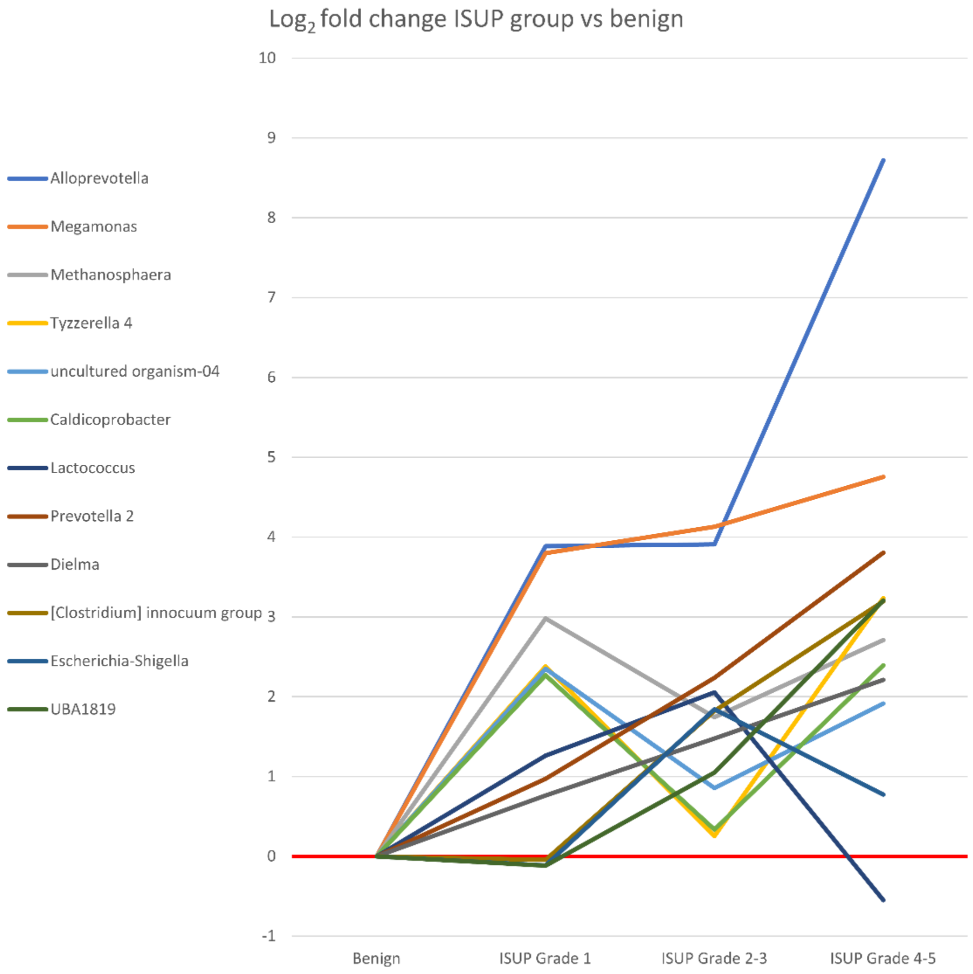

**Figure S6C Log$_2$ fold changes of higher abundance bacteria in prostate cancer according to ISUP grade groups.**

**D**

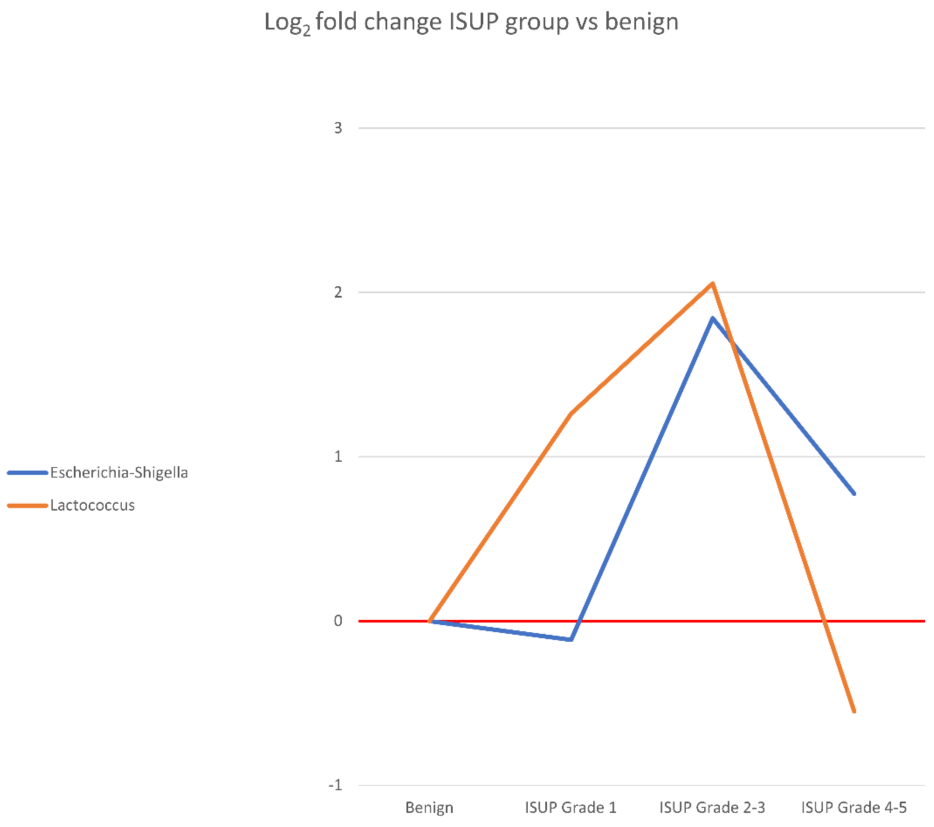

**Figure S6D Log$_2$ fold changes of *Escherichia-Shigella* and *Lactococcus* according to ISUP grade groups.**

#### SUPPLEMENTARY TABLES

##### Table S1 Differential abundance analysis significant results, combined abundance >100, prevalence >10%, FDR-corrected with taxonomy

| Taxonomy | | | | Cancer vs Benign | | | | |
|---|---|---|---|---|---|---|---|---|
| D_0 (Kingdom) D_1 (Phylum) D_2 (Class) D_3 (Order) D_4 (Family) D_5 (Genus) | Combined Abundance | Prevalence in samples (%) | Median of present samples | Max group mean | $Log_2$ fold change | Fold change | P-value | FDR p-value |
| D_0__Archaea, D_1__Euryarchaeota, D_2__Methanobacteria, D_3__Methanobacteriales, D_4__Methanobacteriaceae, D_5__Methanosphaera | 924 | 13 | 6 | 8 | 2.5 | 5.5 | <0.001 | 0.001 |
| D_0__Bacteria, D_1__Actinobacteria, D_2__Actinobacteria, D_3__Corynebacteriales, D_4__Corynebacteriaceae, D_5__Corynebacterium | 1359 | 35 | 4 | 13 | -2.9 | -7.5 | <0.001 | <0.001 |
| D_0__Bacteria, D_1__Bacteroidetes, D_2__Bacteroidia, D_3__Bacteroidales, D_4__Barnesiellaceae, D_5__uncultured-08 | 2665 | 17 | 39 | 20 | 2.1 | 4.2 | 0.001 | 0.009 |
| D_0__Bacteria, D_1__Bacteroidetes, D_2__Bacteroidia, D_3__Bacteroidales, D_4__Marinifilaceae, D_5__Sanguibacteroides | 1124 | 12 | 25 | 9 | 2.1 | 4.2 | 0.001 | 0.009 |
| D_0__Bacteria, D_1__Bacteroidetes, D_2__Bacteroidia, D_3__Bacteroidales, D_4__Muribaculaceae, D_5__CAG-873 | 92781 | 24 | 4 | 678 | 2.3 | 4.8 | 0.001 | 0.01 |
| D_0__Bacteria, D_1__Bacteroidetes, D_2__Bacteroidia, D_3__Bacteroidales, D_4__Muribaculaceae, D_5__uncultured organism-02 | 976 | 13 | 1 | 6 | 2.9 | 7.4 | <0.001 | 0.002 |
| D_0__Bacteria, D_1__Bacteroidetes, D_2__Bacteroidia, D_3__Bacteroidales, D_4__Prevotellaceae, D_5__Alloprevotella | 573293 | 50 | 8 | 4625 | 5.5 | 45.6 | <0.001 | <0.001 |
| D_0__Bacteria, D_1__Bacteroidetes, D_2__Bacteroidia, D_3__Bacteroidales, D_4__Prevotellaceae, D_5__Prevotella 2 | 15028 | 13 | 3 | 136 | 2.2 | 4.4 | 0.001 | 0.02 |
| D_0__Bacteria, D_1__Bacteroidetes, D_2__Bacteroidia, D_3__Bacteroidales, D_4__Prevotellaceae, D_5__Prevotella 9 | 4413629 | 100 | 244 | 30682 | 1.5 | 2.8 | 0.003 | 0.03 |
| D_0__Bacteria, D_1__Bacteroidetes, D_2__Bacteroidia, D_3__Sphingobacteriales, D_4__Lentimicrobiaceae, D_5__uncultured bacterium-09 | 11402 | 24 | 63 | 88 | 2.2 | 4.6 | 0.001 | 0.010 |
| D_0__Bacteria, D_1__Cyanobacteria, D_2__Melainabacteria, D_3__Gastranaerophilales, Ambiguous_taxa, Ambiguous_taxa-07 | 4671 | 16 | 12 | 34 | 2.5 | 5.8 | <0.001 | 0.002 |
| D_0__Bacteria, D_1__Cyanobacteria, D_2__Melainabacteria, D_3__Gastranaerophilales, D_4__gut metagenome, D_5__gut metagenome-02 | 9133 | 11 | 20 | 71 | -2.4 | -5.3 | <0.001 | 0.008 |
| D_0__Bacteria, D_1__Firmicutes, D_2__Bacilli, D_3__Lactobacillales, D_4__Streptococcaceae, D_5__Lactococcus | 2422 | 60 | 5 | 17 | 1.3 | 2.5 | 0.001 | 0.02 |
| D_0__Bacteria, D_1__Firmicutes, D_2__Clostridia, D_3__Clostridiales, D_4__Caldicoprobacteraceae, D_5__Caldicoprobacter | 406 | 23 | 1 | 3 | 1.7 | 3.2 | 0.001 | 0.01 |
| D_0__Bacteria, D_1__Firmicutes, D_2__Clostridia, D_3__Clostridiales, D_4__Christensenellaceae, D_5__Christensenellaceae R-7 group | 171030 | 98 | 175 | 1274 | 1.0 | 2.1 | 0.002 | 0.02 |
| D_0__Bacteria, D_1__Firmicutes, D_2__Clostridia, D_3__Clostridiales, D_4__Clostridiales vadinBB60 group, Ambiguous_taxa-13 | 33714 | 80 | 20 | 295 | 1.6 | 3.1 | <0.001 | 0.002 |
| D_0__Bacteria, D_1__Firmicutes, D_2__Clostridia, D_3__Clostridiales, D_4__Clostridiales vadinBB60 group, D_5__uncultured organism-04 | 518 | 27 | 7 | 4 | 1.6 | 3.0 | 0.001 | 0.01 |
| D_0__Bacteria, D_1__Firmicutes, D_2__Clostridia, D_3__Clostridiales, D_4__Clostridiales vadinBB60 group, D_5__uncultured Thermoanaerobacterales bacterium | 2338 | 43 | 10 | 18 | 1.7 | 3.2 | 0.001 | 0.01 |

| | | | | | | | | |
|---|---|---|---|---|---|---|---|---|
| D_0__Bacteria, D_1__Firmicutes, D_2__Clostridia, D_3__Clostridiales, D_4__Lachnospiraceae, D_5__Butyrivibrio | 204311 | 50 | 4 | 1584 | 2.1 | 4.3 | 0.001 | 0.01 |
| D_0__Bacteria, D_1__Firmicutes, D_2__Clostridia, D_3__Clostridiales, D_4__Lachnospiraceae, D_5__Lachnospiraceae FCS020 group | 7920 | 84 | 17 | 58 | 1.0 | 2.0 | 0.003 | 0.03 |
| D_0__Bacteria, D_1__Firmicutes, D_2__Clostridia, D_3__Clostridiales, D_4__Lachnospiraceae, D_5__Lachnospiraceae UCG-003 | 16360 | 31 | 21 | 106 | 2.4 | 5.4 | <0.001 | 0.003 |
| D_0__Bacteria, D_1__Firmicutes, D_2__Clostridia, D_3__Clostridiales, D_4__Lachnospiraceae, D_5__Moryella | 6991 | 85 | 13 | 61 | -1.5 | -2.7 | <0.001 | 0.001 |
| D_0__Bacteria, D_1__Firmicutes, D_2__Clostridia, D_3__Clostridiales, D_4__Lachnospiraceae, D_5__Tyzzerella 4 | 27590 | 28 | 8 | 237 | 1.8 | 3.5 | 0.004 | 0.03 |
| D_0__Bacteria, D_1__Firmicutes, D_2__Clostridia, D_3__Clostridiales, D_4__Ruminococcaceae, D_5__CAG-352 | 14571 | 19 | 7 | 125 | -3.3 | -9.8 | <0.001 | <0.001 |
| D_0__Bacteria, D_1__Firmicutes, D_2__Clostridia, D_3__Clostridiales, D_4__Ruminococcaceae, D_5__Hydrogenoanaerobacterium | 2547 | 43 | 7 | 22 | 2.0 | 4.0 | <0.001 | 0.001 |
| D_0__Bacteria, D_1__Firmicutes, D_2__Clostridia, D_3__Clostridiales, D_4__Ruminococcaceae, D_5__Oscillospira | 1155 | 22 | 4 | 7 | 1.5 | 2.8 | 0.006 | 0.05 |
| D_0__Bacteria, D_1__Firmicutes, D_2__Clostridia, D_3__Clostridiales, D_4__Ruminococcaceae, D_5__UBA1819 | 27061 | 92 | 18 | 193 | 1.1 | 2.2 | 0.005 | 0.04 |
| D_0__Bacteria, D_1__Firmicutes, D_2__Erysipelotrichia, D_3__Erysipelotrichales, D_4__Erysipelotrichaceae, D_5__[Clostridium] innocuum group | 466 | 23 | 3 | 4 | 2.1 | 4.2 | <0.001 | 0.002 |
| D_0__Bacteria, D_1__Firmicutes, D_2__Erysipelotrichia, D_3__Erysipelotrichales, D_4__Erysipelotrichaceae, D_5__Coprobacillus | 1284 | 17 | 6 | 10 | 4.4 | 20.7 | <0.001 | <0.001 |
| D_0__Bacteria, D_1__Firmicutes, D_2__Erysipelotrichia, D_3__Erysipelotrichales, D_4__Erysipelotrichaceae, D_5__Dielma | 167 | 21 | 3 | 1 | 1.5 | 2.9 | 0.001 | 0.01 |
| D_0__Bacteria, D_1__Firmicutes, D_2__Negativicutes, D_3__Selenomonadales, D_4__Acidaminococcaceae, D_5__Acidaminococcus | 112181 | 43 | 7 | 903 | 2.5 | 5.6 | <0.001 | 0.005 |
| D_0__Bacteria, D_1__Firmicutes, D_2__Negativicutes, D_3__Selenomonadales, D_4__Veillonellaceae, D_5__Anaeroglobus | 12425 | 44 | 7 | 141 | -3.0 | -7.8 | <0.001 | <0.001 |
| D_0__Bacteria, D_1__Lentisphaerae, D_2__Lentisphaeria, D_3__Victivallales, D_4__vadinBE97, D_5__uncultured rumen bacterium-04 | 528 | 22 | 8 | 4 | 1.5 | 2.8 | 0.004 | 0.04 |
| D_0__Bacteria, D_1__Proteobacteria, D_2__Gammaproteobacteria, D_3__Betaproteobacteriales, D_4__Burkholderiaceae, D_5__Alcaligenes | 700 | 54 | 3 | 4 | -1.2 | -2.2 | 0.003 | 0.03 |
| D_0__Bacteria, D_1__Proteobacteria, D_2__Gammaproteobacteria, D_3__Betaproteobacteriales, D_4__Burkholderiaceae, D_5__Parasutterella | 77911 | 90 | 44 | 495 | -1.3 | -2.5 | 0.002 | 0.02 |
| D_0__Bacteria, D_1__Proteobacteria, D_2__Gammaproteobacteria, D_3__Betaproteobacteriales, D_4__Neisseriaceae, D_5__uncultured-29 | 1206 | 12 | 17 | 10 | 2.5 | 5.5 | <0.001 | 0.002 |
| D_0__Bacteria, D_1__Proteobacteria, D_2__Gammaproteobacteria, D_3__Enterobacteriales, D_4__Enterobacteriaceae, Ambiguous_taxa-29 | 139 | 27 | 2 | 1 | 1.3 | 2.4 | 0.003 | 0.03 |
| D_0__Bacteria, D_1__Proteobacteria, D_2__Gammaproteobacteria, D_3__Enterobacteriales, D_4__Enterobacteriaceae, D_5__Citrobacter | 533 | 13 | 7 | 4 | 1.7 | 3.4 | 0.002 | 0.02 |
| D_0__Bacteria, D_1__Proteobacteria, D_2__Gammaproteobacteria, D_3__Enterobacteriales, D_4__Enterobacteriaceae, D_5__Escherichia-Shigella | 551481 | 98 | 229 | 3314 | 1.3 | 2.5 | 0.003 | 0.03 |
| D_0__Bacteria, D_1__Synergistetes, D_2__Synergistia, D_3__Synergistales, D_4__Synergistaceae, D_5__Jonquetella | 27709 | 40 | 14 | 293 | -2.4 | -5.2 | <0.001 | 0.001 |
| D_0__Bacteria, D_1__Verrucomicrobia, D_2__Verrucomicrobiae, D_3__Opitutales, D_4__Puniceicoccaceae, D_5__uncultured-32 | 4177 | 77 | 10 | 35 | 2.4 | 5.3 | <0.001 | 0.002 |

**Table S2 PICRUSt results P<0.10**

| KEGG Pathway | Benign | | Cancer | | p-value |
|---|---|---|---|---|---|
| | Median | (IQR) | Median | (IQR) | |
| Mineral absorption | 2522 | (1197-8072) | 4044 | (1868-18695) | 0,008 |
| Steroid hormone biosynthesis | 8457 | (3373-16084) | 11717 | (4937-21186) | 0,022 |
| Retinol metabolism | 21605 | (8832-29630) | 23174 | (11812-35315) | 0,042 |
| Bladder cancer | 37 | (11-247) | 171 | (13-635) | 0,051 |
| Bacterial invasion of epithelial cells | 49 | (11-256) | 119 | (23-673) | 0,053 |
| African trypanosomiasis | 426 | (165-860) | 591 | (248-1252) | 0,057 |
| Fluorobenzoate degradation | 131 | (27-395) | 263 | (50-821) | 0,067 |
| Arachidonic acid metabolism | 37003 | (14831-69756) | 51649 | (19471-80097) | 0,073 |
| Carbohydrate digestion and absorption | 15218 | (5621-27855) | 21879 | (8317-33654) | 0,074 |
| Biosynthesis of siderophore group nonribosomal peptides | 30806 | (12789-35865) | 32048 | (24926-40301) | 0,083 |
| Glycan biosynthesis | 28853 | (15104-39625) | 33540 | (19369-41826) | 0,083 |
| Chagas disease (American trypanosomiasis) | 231 | (70-509) | 317 | (98-870) | 0,086 |
| Isoquinolone alcaloid biosynthesis | 59184 | (45862-71701) | 62282 | (45863-80307) | 0,096 |
